# PerTexP: scenario-based exploration of pertussis dynamics under maternal and infant vaccination

**DOI:** 10.64898/2026.03.05.26347721

**Authors:** Anna Autoriello, Sabrina Averga, Bruno Buonomo, Rossella Della Marca, Alfredo Guarino, Cristina Moracas, Emanuela Penitente, Marco Poeta

## Abstract

We introduce `PerTexP` (Pertussis Time Exploration), an interactive modelling tool designed to investigate pertussis transmission dynamics and to support the evaluation of vaccination strategies and short-term projections. `PerTexP` allows users to explore and compare maternal, infant, and non–infant booster vaccination scenarios and to assess their potential impact on disease transmission, with a particular focus on the Italian epidemiological context. The tool is based on a discrete-time, stage–structured compartmental model with two age classes. By enabling rapid scenario-based analyses, `PerTexP` supports evidence-informed decision-making and provides transparent insights into how alternative vaccination strategies may shape pertussis dynamics. Combining accessibility, flexibility, and methodological rigour, `PerTexP` offers a practical resource for researchers and public health practitioners interested in exploring and comparing pertussis control strategies.

## 1 Introduction

Despite the widespread availability of effective vaccines, pertussis remains a significant public health concern in many high-income countries. Over the past decades, several regions have experienced a resurgence of reported pertussis cases, characterised by recurrent outbreaks and an overall increase in incidence [6]. This re-emergence has been observed even in settings with high vaccination coverage, raising important questions about the mechanisms sustaining transmission [30]. In particular, the growing incidence among adolescents and adults, who often experience mild or atypical symptoms, represents a critical challenge for pertussis control, as these individuals may act as unrecognised sources of infection for vulnerable infants [9, 52]. This evidence highlights the need for effective intervention strategies that go beyond childhood vaccination alone and account for waning immunity and heterogeneous vaccine uptake across age groups.

Alongside clinical and public health measures, quantitative modelling approaches can provide valuable support for decision-making by offering a systematic framework to explore transmission dynamics, evaluate vaccination strategies, and assess their potential impact over short- and medium-term horizons [4, 17]. In this context, there is a clear need for modelling tools that are both mathematically rigorous and accessible to practitioners, enabling the exploration of alternative scenarios and supporting evidence-based planning. Motivated by these considerations, this work aims to develop `PerTexP` (Pertussis Time Exploration), a user-friendly MATLAB-based modelling tool [57] for investigating pertussis transmission dynamics and vaccination strategies. We also provide the epidemiological and vaccination background and review the mathematical modelling literature underpinning its development. The tool supports short-term projections, the comparison of different vaccination strategies, and the assessment of intervention effectiveness, with particular attention to the Italian epidemiological context. The vaccination scenarios considered in `PerTexP` include maternal immunisation, infant primary vaccination, and booster interventions. To place the scope and practical relevance of `PerTexP` in context, we first briefly summarise the main epidemiological and clinical features of pertussis.

Pertussis, also known as whooping cough, is a highly contagious respiratory infection caused by the bacterium Bordetella pertussis. The disease spreads from person to person through respiratory droplets, and individuals can transmit the bacteria from the onset of symptoms for at least two weeks after coughing begins [7]. The incubation period, approximately corresponding to the latent stage, typically ranges from 7 to 10 days, and the overall disease course can last 2–3 months [35]. Pertussis poses a particular threat to neonates and infants, representing a significant cause of morbidity and mortality in this age group. Globally, more than 151 000 pertussis cases were reported in 2018, underscoring the ongoing public health relevance of the disease [68]. In Europe, over 25 000 cases were reported in 2023, with an additional 32 000 cases recorded between January and March 2024. In 2024, a major outbreak was detected in Italy with 108 hospitalisations among neonates and younger infants and 3 deaths [45]; in the same context, national surveillance data reported an incidence of 9.9 cases per million inhabitants from January to May 2024 [37].

Clinical manifestations of pertussis are especially severe in infants under one year of age. Common complications include cough–induced haemorrhagic events and respiratory infections such as pneumonia and bronchopneumonia. The most severe forms involve the central nervous system, with pertussis-associated encephalopathy resulting from both hypoxia during paroxysmal coughing and the direct action of pertussis toxin. In Italy, mortality due to pertussis has drastically decreased over the past century, from approximately 10 per thousand in the early 20th century to 0.01 per thousand in recent years, although it can still reach 0.5-1 per thousand in infants under one year of age [38].

Current control measures primarily focus on vaccination-based interventions designed to reduce transmission and provide early-life protection. A major approach includes maternal vaccination during pregnancy. The Tdap (tetanus, diphtheria, acellular pertussis) vaccination is recommended for pregnant women starting from 16 weeks of gestation, typically administered around the mid-pregnancy scan at 20 weeks. For optimal protection of the newborn, it is best administered before 32 weeks. Maternal Tdap vaccination confers passive immunity to the infant, significantly reducing the risk of pertussis in early life [40].

An evaluation by the Centers for Disease Control and Prevention (CDC) reported that Tdap administration during the third trimester prevents 78% of pertussis cases and 91% of pertussis-related hospitalisations in infants younger than 2 months [53]. Accordingly, missed maternal vaccination can translate into substantial infant morbidity: during the 2024 outbreak in Italy, infants born to unvaccinated mothers were disproportionately affected, and hospitalisation rates were higher in regions with lower maternal vaccination coverage [46]. The protection lasts until approximately 8 weeks of age, when infants begin their own pertussis vaccination schedule [8].

Another key strategy is childhood vaccination. Typically, the vaccination schedule includes a three-dose primary series in the first year of life (starting around 8 weeks of age), followed by booster doses in early childhood and later in adolescence and adulthood to maintain vaccine–induced protection [8, 29]. These booster doses are necessary because protection after pertussis vaccination wanes over time: the DTaP vaccine (diphtheria, tetanus, acellular pertussis), widely used in high-income countries, shows an efficacy of approximately 98% within the first year following the final dose of the primary series, declining to around 71% after five years [34]. Evidence suggests that immunity decreases substantially between 4 and 7 years after vaccination, underscoring the need for timely booster doses during childhood and adolescence [51]. Among adolescents, Tdap provides about 73% protection in the first year after vaccination, dropping to approximately 34% by four years post-vaccination [1]. Compared to the currently used acellular vaccine, the older whole–cell pertussis vaccine confers longer-lasting protection, but is associated with a higher risk of adverse events and is now rarely used in high–income settings [54, 64].

Mathematical models provide a complementary framework for understanding the transmission dynamics of infectious diseases and assessing the potential impact of intervention strategies [22]. In particular, models based on dynamical systems theory are valuable in contexts where epidemiological data are limited, incomplete, or affected by substantial uncertainty, as they allow the integration of biological assumptions with available empirical evidence and enable the exploration of plausible scenarios and qualitative system behaviour [31, 41]. In the case of pertussis, limitations in data quality and completeness may hinder reliable quantitative inference [11], making mathematical models a useful tool for investigating transmission mechanisms and evaluating intervention strategies at a qualitative level.

Some modelling studies have investigated pertussis transmission using continuous-time compartmental models, often incorporating age structure and vaccination. Early contributions by Hethcote and coworkers laid the foundations for age–structured modelling of pertussis dynamics. In particular, a highly detailed compartmental model with multiple age classes and epidemiological states is introduced in [20], together with parameter estimation based on available data. Subsequent work focuses on the analytical and numerical exploration of simplified versions of this model [21, 24], as well as on the use of computer simulations to assess the potential impact of booster vaccination strategies in specific settings, such as Australia [23].

Within the context of evaluating vaccination strategies against pertussis, van Rie et al. [62] employed an age–structured deterministic compartmental model to prospectively compare alternative adolescent and adult immunisation policies. Their analysis accounted for both the direct protection of vaccinated individuals and the indirect effects on transmission, highlighting possible benefits for reducing the burden of severe pertussis in young infants.

Motivated by the pertussis epidemic observed in the Netherlands in 1996–1997, van Boven et al. [59] developed an age–structured SIR (Susceptible–Infected–Recovered) model to investigate the combined effects of waning immunity and under–reporting. By distinguishing between primary infections in immunologically naïve individuals and reinfections in previously immunised individuals, the study emphasised the potential role of unobserved infections in sustaining transmission. In line with the epidemiological context of the time, the model assumes that all individuals are born susceptible and does not include maternal vaccination.

The role of waning immunity was further explored by Wearing et al. [65], who extended the classical SEIR (Susceptible–Exposed–Infected–Recovered) framework to include reinfection, vaccination, and immune boosting. By analysing both deterministic and stochastic formulations, their work examined how different assumptions on immunity loss influence epidemic persistence, inter-epidemic periods, and herd immunity, again highlighting the epidemiological relevance of subclinical infections and reinfections. More recently, Tian et al. [58] proposed an age–structured SIR model with vaccinations in which the infectious class is explicitly partitioned into dominant (symptomatic) and covert (subclinical) infections. This approach allows for a more explicit representation of hidden transmission within a vaccination framework, further underscoring the importance of unobserved infections in pertussis dynamics.

The tool presented here, `PerTexP`, is based on a discrete–time compartmental model, which is well suited to short-term projections and the comparison of vaccination scenarios over discrete reporting intervals, particularly in the presence of relatively low and variable pertussis case counts. The modelling approach is inspired by the stage–structured discrete epidemic model introduced by Li and Wang [33], who proposed a SIS model for a generic infection distinguishing immature and mature individuals. Building on this perspective, we consider a stage–structured compartmental model with two age classes: *infants*, comprising individuals aged 0 to 11 months, and *non–infants*, comprising individuals aged 1 year and older. This structure reflects age–specific differences in pertussis severity, mortality risk, and vaccination schedule, since infants under 1 year have a higher risk of hospitalisation and disease–induced mortality, and the primary vaccination series is completed before 1 year of age [7]. The model explicitly incorporates maternal vaccination, routine infant immunisation, and booster vaccination in the non–infant age group. We conduct a qualitative analysis of the model and derive the control reproduction number, which provides a threshold condition for the sustained circulation of pertussis in the population. Applied to epidemiological scenarios concerning the Italian context, `PerTexP` enables the assessment of the impact of maternal immunisation, infant primary vaccination, and non–infant booster uptake, supporting the systematic evaluation of vaccination strategies under realistic assumptions.

The rest of the paper is organised as follows. Section 2 presents the model embedded within `PerTexP`, Section 3 discusses the reproduction numbers, Section 4 describes the model parametrisation, Section 5 illustrates the main outcomes of `PerTexP` and Section 6 concludes the paper with a discussion of the results and perspectives for future work.

## 2 The model embedded within PerTexP

### 2.1 Age–group structure and compartments

`PerTexP` is built upon a discrete–time compartmental model with the two age classes defined in Section 1: *infants* (from 0 to 11 months) and *non–infants* (1 year or older). We model the discrete time with a weekly step, so that *t* denotes the week. The infant and non–infant population sizes at time step *t* are denoted by *N*_1_(*t*) and *N*_2_(*t*), respectively. Within each age group, pertussis transmission is modelled using a SIR–type compartmental model, where individuals are classified as susceptible *S*_*i*_(*t*), infected *I*_*i*_(*t*) and recovered *R*_*i*_(*t*), *i* = 1, 2. This formulation assumes that infected individuals are immediately infectious and therefore neglects the latency period, during which the individual has been exposed to the disease but is not yet infectious. However, the latency period of pertussis (typically ranging from 7 to 10 days) is relatively short compared to the infectious period, which can last for several weeks [7, 35]. For this reason, we do not include the exposed compartment in `PerTexP`, as the brief latency is unlikely to significantly influence the overall epidemic dynamics. While more complex models incorporating additional epidemiological features have been proposed [20, 21, 23, 65], they require the estimation of a larger number of parameters, leading to a greater degree of uncertainty in model outcomes [49, 50, 55]. Given the limited availability of surveillance data, adopting a SIR model within each age group supports a more robust quantitative scenario comparison while capturing the key features of the underlying dynamics. In each age group, we consider vaccination strategies by including a compartment for vaccinated individuals. Regarding vaccination of infants under 1 year of age, two strategies are currently available in Italy, which is the reference setting for this study: maternal vaccination, which protects the newborns during the first two months of life; and primary vaccination of infants. The DTaP primary vaccination series against pertussis is included in the national immunisation schedule and consists of three doses: the first at 2 months of age, the second at 4 months, and the third at 10 months [8,29]. To balance the biological features with model simplicity, we represent infant vaccination through a single compartment *V*_1_ of immunised individuals. This compartment includes those protected by maternal antibodies and/or who have received at least two doses of the DTaP vaccine. Conversely, infants who are not protected at birth or have received only one dose (and are therefore considered not protected) are included in the susceptible compartment *S*_1_. Regarding immunisation of non–infants, the vaccination schedule in Italy includes a single booster dose every 10 years [8]. For this reason, we incorporated a compartment *V*_2_ of immunised individuals, which includes both non–infants who have protection due to the booster dose and infants who were immunised in *V*_1_ and, upon ageing into the non–infant class, move to *V*_2_ while maintaining protection. Both vaccinations are not effective in all subjects, and we account for this by introducing an ineffectiveness factor *σ*_*i*_ ∈ (0, 1), *i* = 1, 2, representing the fraction of vaccinated individuals who fail to develop protective immunity. Both vaccinations are also temporary, losing their effectiveness after approximately 10 years [8, 35]. After this period, vaccinated individuals become susceptible again. Since the compartment *V*_1_ collects two types of protected infants (through either maternal antibodies or primary vaccination series), we neglect any transition from *V*_1_ to *S*_1_ for two reasons: (*i*) the duration of the immunity provided by the primary DTaP series is much longer than the timespan of the infant age class; (*ii*) although maternally derived antibodies wane within approximately two months after birth, we assume that these infants promptly receive the first DTaP dose and that the combined protection from maternal antibodies and the first vaccine dose remains sufficiently high to keep them effectively protected throughout the time spent in the infant age class. Accordingly, waning of vaccine–induced immunity is considered only among non–infants, and vaccinated individuals in *V*_2_ return to *S*_2_ with probability *ω* at each time step.

Let *S*_*i*_(*t*), *V*_*i*_(*t*), *I*_*i*_(*t*), and *R*_*i*_(*t*) denote the numbers of susceptible, protected, infectious and recovered individuals in the *i*–th age group, *i* = 1, 2, at discrete time step *t*. The total population in each age group is given by

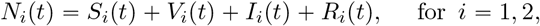

and the total population size is *N* (*t*) = *N*_1_(*t*) + *N*_2_(*t*).

### 2.2 Demography, ageing and deaths

At each time step, individuals may move from the first to the second age group with probability *η*. Their health status is assumed to remain unchanged during the transition from one age group to another. The model incorporates demographic turnover, a process that is essential for modelling prenatal vaccination. It is represented by a constant inflow denoted by Λ that enters the first age class, and by natural mortality, which occurs with probability *µ*_*i*_ per time step in the *i*–th age group. When writing the equations of the model embedded in `PerTexP`, we use for convenience the survival probability *r*_*i*_, *i* = 1, 2, which can be trivially computed from the natural death probability as *r*_*i*_ = 1 − *µ*_*i*_. In addition to natural mortality, `PerTexP` includes disease–induced death in the first age group, occurring with probability *d*. As a result, survival in infants depends on health status: uninfected infants survive with probability *r*_1_, while infected infants have a reduced survival probability, denoted as *r*_3_ = 1 − (*µ*_1_ + *d*). Disease–induced mortality is not considered in the second group, since the risk of death from pertussis strongly declines after the first year of life [36].

Under these assumptions, the population sizes in the two age groups evolve according to a baseline demographic subsystem given by the following system of difference equations. This recurrence updates the state variables at discrete time *t* +1 based on the system state at time *t* and on the events occurring in the interval [*t, t* + 1):

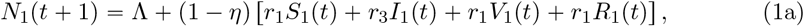

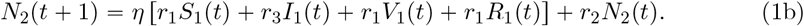

In particular, equation (1a) computes the variable *N*_1_(*t* + 1) as the sum of two contributions: the weekly recruitment of newborns, equal to Λ, and the infants who were already in *N*_1_ at time *t* that have survived the week and have not yet moved to the non-infant group. Given that the weekly ageing probability is equal to *η* and the survival probabilities for infants can be *r*_1_ or *r*_3_ depending on their health status, the number of infants who remain in *N*_1_ at time *t* + 1 is equal to (1 − *η*)[*r*_1_*S*_1_(*t*) + *r*_3_*I*_1_(*t*) + *r*_1_*V*_1_(*t*) + *r*_1_*R*_1_(*t*)].

Similarly, equation (1b) computes the variable *N*_2_(*t* + 1) by adding the infants who survive during the week and age, moving into the non–infant class, to the individuals who were already non–infants at time *t* and remain in the same age class, surviving with probability *r*_2_. The inflow term given by the infants who age is given by *η* [*r*_1_*S*_1_(*t*) + *r*_3_*I*_1_(*t*) + *r*_1_*V*_1_(*t*) + *r*_1_*R*_1_(*t*)].

### 2.3 Transmission and vaccination dynamics

A fraction *p* of newborns is assumed to be protected via maternal antibodies and is thus allocated in the protected compartment *V*_1_. The remaining proportion, 1 − *p*, enters the susceptible compartment *S*_1_. At each time step, susceptible individuals in the *i*– th age group may either become infected following exposure to the pathogen or receive vaccination, after which they move to the vaccinated compartment *V*_*i*_. A fraction *ψ*_*i*_ of susceptible individuals is vaccinated at each time step. Transmission of pertussis occurs through contact between susceptible (or vaccinated) individuals and infectious individuals in either age group. The risk of infection is modelled via the *contagion probability function G*_*i*_(·), which represents the probability that a susceptible or vaccinated individual in group *i* becomes infected during the time interval [*t, t* + 1). Vaccinated individuals experience a reduced risk of infection, expressed by the term *σ*_*i*_*G*_*i*_. Consequently, the probability of escaping infection in a given time step is 1−*G*_*i*_ for susceptibles and 1−*σ*_*i*_*G*_*i*_ for vaccinated individuals. We model transmission using a *frequency–dependent* formulation, that is, the per–susceptible risk of infection due to infectious individuals of the *j*–th age group scales with the prevalence *I*_*j*_{*N*_*j*_. Since pertussis transmission occurs through contact between susceptible and infectious individuals across all age groups, we assume that *G*_*i*_ = *G*_*i*_(*λ*_*i*_),

Where

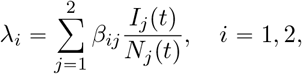

where the age–specific transmission rate *β*_*ij*_ = *c*_*ij*_*π*_*ij*_ is defined as the product of the average number of contacts per time step *c*_*ij*_ between individuals in age groups *i* and *j*, and the probability *π*_*ij*_ of successful transmission given contact with an infectious individual from group *j*. The contact structure is represented by a *contact matrix* 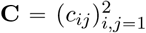, where each element *c*_*ij*_ denotes the average number of contacts per time step that an individual in age group *i* has with individuals in age group *j*. This frequency–dependent approach is well–suited to pathogens that spread over heterogeneous contact structures [31] and is consistent with the one adopted for continuous age–structured pertussis models [20], as well as for discrete–time models of other airborne diseases that require age–structure, since these primarily affect infants [47]. The functional form of *G*_*i*_ is given by:

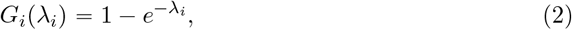

consistent with standard assumptions on contagion probabilities and with Poisson infection processes. A more detailed discussion of the rationale behind this choice, together with the mathematical properties that *G*_*i*_ is required to satisfy, is provided in the Supplementary Material, § S1.

At each time step *t*, infected individuals in class *I*_*i*_ recover with probability *γ*_*i*_ and move to the recovered compartment *R*_*i*_, or stay infectious with probability 1 −*γ*_*i*_, for *i* = 1, 2. In addition, recovered individuals in the non–infant group may lose natural immunity with probability *ν* and return to the susceptible class *S*_2_ in the next time step; otherwise, they remain in *R*_2_ with probability 1 −*ν*. Similarly, vaccinated non–infant individuals may lose vaccine–induced protection with probability *ω*, leading to a return to the susceptible class *S*_2_ at the next time step.

### 2.4 The balance equations

The processes governing the mathematical model underlying `PerTexP`, illustrated in the flowchart in Figure 1, are mathematically described through a discrete–time system. The time variable *t* denotes the weeks, so the nonnegative integer *t* denotes the current week and *t* + 1 the next one. At each step, the state of the system is updated by accounting for the relevant processes: recruitment, infection, vaccination, recovery, ageing, mortality and waning of protection. The resulting equations are *balance equations*, since each state variable at time *t*+1 is obtained as the balance between those who were in the compartment at week *t* and remain there after surviving the week, plus inflows from other compartments, minus the corresponding outflows. The model is given by:

**Figure 1:**
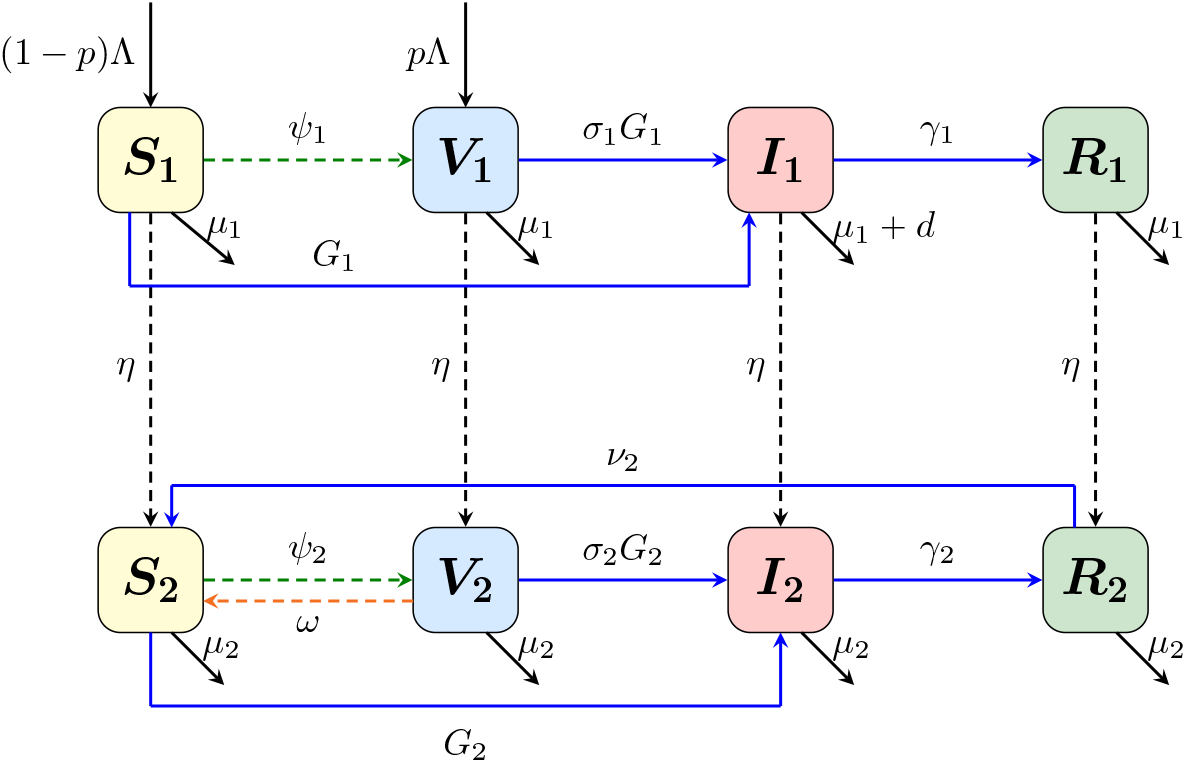
Flowchart of the discrete–time model underlying the MATLAB tool `PerTexP`. Blue arrows indicate infection, recovery, and waning of natural immunity. Black arrows denote demographic processes (recruitment, ageing and deaths), while orange arrows denote waning of vaccine-induced protection.

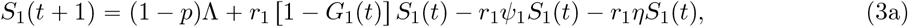

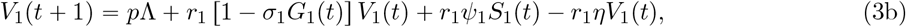

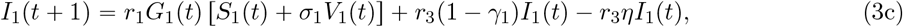

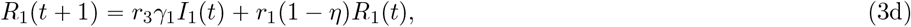

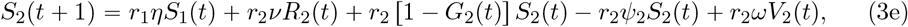

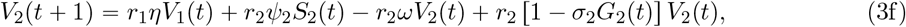

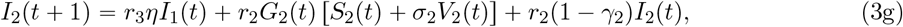

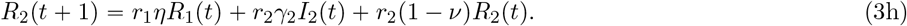

The initial conditions, representing the state of the system at the beginning of the observation period, satisfy

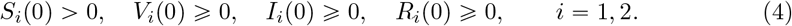

The basic mathematical properties of the discrete–time model (i.e., positivity of the solutions and biologically feasible region) are reported in the Supplementary Material, § 2.

## 3 The reproduction numbers

One of the most commonly used indicators to quantify the transmission potential of a pathogen is the *basic reproduction number*, ℛ_0_, defined as the expected number of secondary infections generated by a source case introduced into a wholly susceptible population, during its entire period of infectiousness and in the absence of pre–existing immunity or interventions [13, 19]. When specific mitigation measures (e.g., vaccination or contact reduction) are in place, the corresponding quantity is the *control reproduction number*, usually denoted by ℛ_*c*_ [18]. By construction, ℛ_*c*_ < ℛ_0_. One of the most useful properties of ℛ_0_ (or ℛ_*c*_, when interventions are in place) is its role as an epidemic threshold: typically, if ℛ_0_ < 1, each infected individual generates, on average, less than one secondary case and the disease is expected to die out. Conversely, if ℛ_0_ > 1, the pathogen can invade a wholly susceptible population and spread. Estimates of these reproduction numbers can therefore provide valuable insights when designing and comparing control strategies during outbreaks of infectious diseases [12, 19, 60].

We compute ℛ_0_ and ℛ_*c*_ in terms of the model parameters using the so–called *next–generation matrix* (NGM) approach, initially developed for continuous–time epidemic models [13, 61] and later extended to the discrete–time setting [2]. This approach summarises the transmission processes through a square matrix, **NGM**, whose dimension is equal to the number of infected compartments. In our model, new infections occur in the two infectious compartments, *I*_1_ and *I*_2_, so **NGM** is a 2 × 2 matrix. In the discrete–time setting, the next–generation matrix is defined as

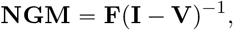

where **F** is the *fertility* matrix, **V** is the *transition* matrix, and **I** denotes the identity matrix. The matrices **F** and **V** are defined, respectively, as the Jacobian matrix of the new–infection terms and the Jacobian matrix of the transition terms (i.e., age progression, recovery or death), among infected compartments, both evaluated at the *disease–free equilibrium*, that is, the steady–state where there are no infected individuals. The control reproduction number is then the spectral radius of **NGM**, i.e., the dominant eigenvalue. Beyond this mathematical definition, the next–generation matrix also admits an epidemiological interpretation: its (*i, j*)–entry represents the expected number of secondary cases in compartment *i* produced by a single infectious individual in compartment *j*.

We first compute the expression of the control number ℛ_*c*_:

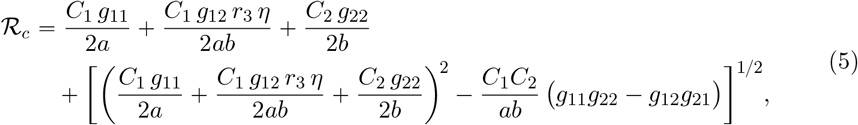

where *a* = 1 − *r*_3_(1 − *γ*_1_ − *η*), *b* = 1 − *r*_2_(1 − *γ*_2_), and

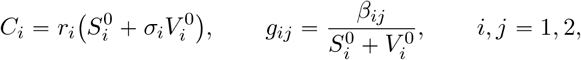

while 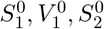 and 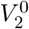 denote the disease–free equilibrium coordinates. For the sake of brevity, the explicit expressions of the disease–free equilibrium coordinates are reported in the Supplementary Material, § 3. The expression of the basic reproduction number can be recovered from that of ℛ_*c*_ by setting the vaccination–related parameters equal to zero, that is, *ψ*_1_ = *ψ*_2_ = 0 and *p* = 0. This yields the following expression of ℛ_0_:

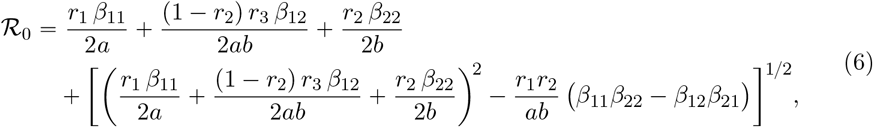

where the parameters *a* and *b* are defined as in the expression of ℛ_*c*_. A more detailed derivation of the reproduction numbers, together with the interpretation of the next–generation matrix, is given in the Supplementary Material, § 3.

## 4 Parametrisation

The parameters of `PerTexP`, together with their baseline values, the simulation time horizon and the initial conditions are summarised in Table 2. The baseline values are informed by official 2024 Italian demographic statistics [26] and estimates from the clinical and epidemiological literature. The remainder of this section details the parametrisation.

**Table 1:**
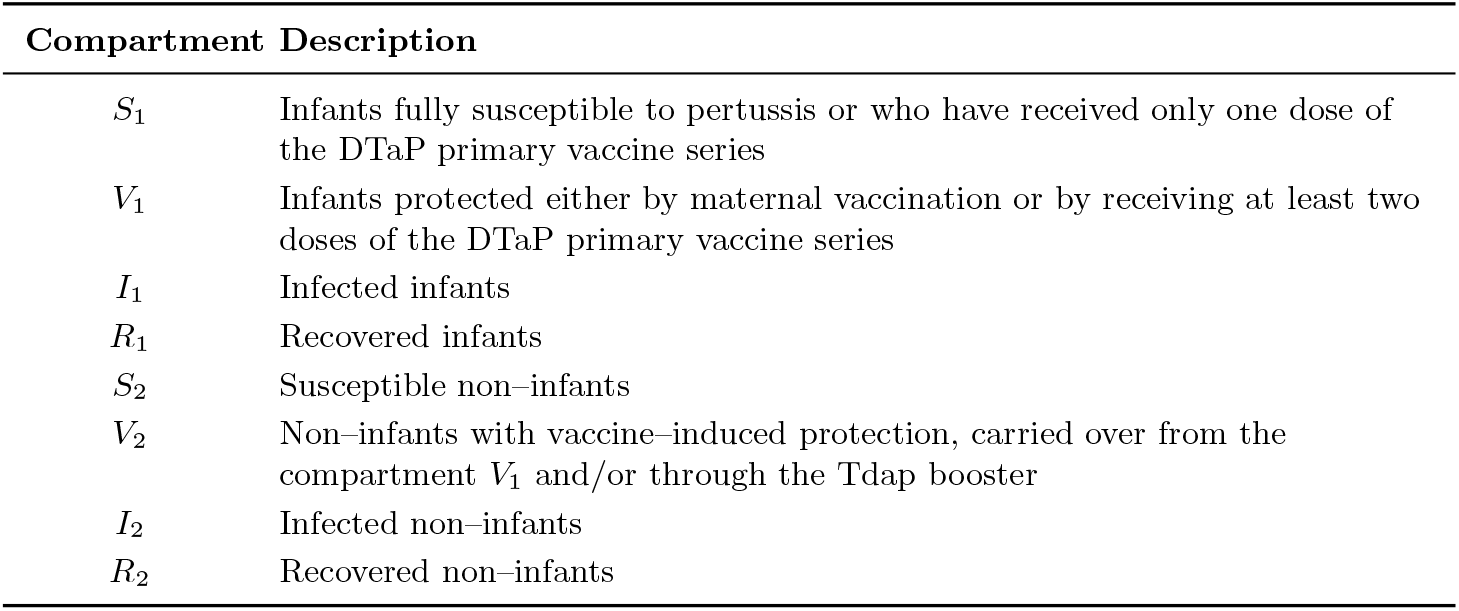
Description of the PerTexP epidemiological compartments.

**Table 2:** Description, baseline values and sources for PerTexP parameters, based on official 2024 Italian demographic statistics and estimates drawn from clinical and epidemiological literature.

| Parameter | Description | Baseline value | Source |
| --- | --- | --- | --- |
| $t_f$ | Temporal horizon | 260 weeks | Assumed |
| $\tilde{N}_1$ | Infant population at the DFE | 380 630 individuals | [26] |
| $\tilde{N}_2$ | Non-infants population at the DFE | 58 590 600 individuals | [26] |
| $S_1(0)$ | Initial number of susceptibles infants | $6.3001 \cdot 10^4$ | See (13) |
| $V_1(0)$ | Initial number of vaccinated infants | $3.1762 \cdot 10^5$ | Assumed |
| $I_1(0)$ | Initial number of infected infants | 5 | Assumed |
| $R_1(0)$ | Initial number of recovered infants | 0 | Assumed |
| $S_2(0)$ | Initial number of susceptibles non-infants | $5.427 \cdot 10^7$ | See (13) |
| $I_2(0)$ | Initial number of infected non-infants | 15 | Assumed |
| $V_2(0)$ | Initial number of vaccinated non-infants | $4.321 \cdot 10^6$ | Assumed |
| $R_2(0)$ | Initial number of recovered non-infants | 4064 | Assumed |
| $\Lambda$ | Weekly recruitment of newborns | 7 338 individuals | See (10) |
| $p$ | Fraction of infants with immunity given by maternal vaccination | 0.626 | [46] |
| $\sigma_1$ | DTaP vaccine ineffectiveness factor | 0.170 | [48] |
| $\sigma_2$ | Tdap vaccine ineffectiveness factor | 0.080 | [63] |
| $\psi_1$ | Weekly probability of vaccination for infants | $5.492 \cdot 10^{-2}$ | [27] |
| $\psi_2$ | Weekly probability of non-infant vaccination | $3.176 \cdot 10^{-5}$ | [32] |
| $\omega$ | Weekly probability of losing vaccination-induced immunity (non-infants) | $1.921 \cdot 10^{-3}$ | [51] |
| $\mu_{1,\text{ann}}$ | Annual probability of death for infants | $2.572 \cdot 10^{-3}$ | [25] |
| $\mu_{2,\text{ann}}$ | Annual probability of death for non-infants | $6.413 \cdot 10^{-3}$ | See (11) |
| $\rho_{\text{cfr}}$ | Probability that an infected infant dies from pertussis before leaving the infectious state | $6.300 \cdot 10^{-3}$ | [6] |
| $\eta$ | Weekly probability of ageing from infants to non-infants | $1.905 \cdot 10^{-2}$ | See (12) |
| $\beta_{11}$ | Transmission rate from infants to infants | $0.81 \text{ week}^{-1}$ | Assumed |
| $\beta_{12}$ | Transmission rate from non-infants to infants | $3.0 \text{ week}^{-1}$ | Assumed |
| $\beta_{21}$ | Transmission rate from infants to non-infants | $0.002 \text{ week}^{-1}$ | Assumed |
| $\beta_{22}$ | Transmission rate from non-infants to non-infants | $0.30 \text{ week}^{-1}$ | Assumed |
| $\gamma_1$ | Weekly probability of recovery for infants | $2.835 \cdot 10^{-1}$ | [10] |
| $\gamma_2$ | Weekly probability of recovery for non-infants | $2.835 \cdot 10^{-1}$ | [10] |
| $\nu$ | Weekly probability of losing infection-acquired immunity (non-infants) | $1.921 \cdot 10^{-3}$ | [66] |

### 4.1 Demographic parameters

To capture multiple consecutive epidemic seasons, we consider a five–year time horizon. The time step is one week, so the final time is *t*_*f*_ = 260 weeks. Over this relatively short horizon, the total sizes of the two age groups, N_1_(t) and *N*_2_(*t*), are held at their demographic steady–state values, 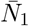 and 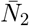. These correspond to the unique equilibrium of the disease–free demographic subsystem,

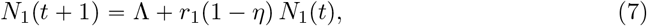

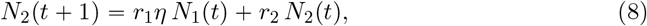

determined by the balance between recruitment, natural mortality, and ageing. We set 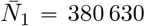 and 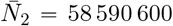, matching the numbers of individuals aged less than one year and those aged one year or more in Italy in January 2024 [26].

The value of the weekly survival probability for infants, *r*_1_, is derived from the annual probability of death for children under 1 year reported by the Italian National Institute of Statistics (ISTAT), 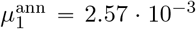 (i.e. 257 deaths per 100 000 infants per year). Using the standard discrete–time conversion [42],

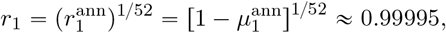

where 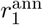 denotes the annual survival probability.

The weekly survival probability for infected infants, *r*_3_, which also incorporates the weekly probability of disease–induced death *d*, is calibrated using the case fatality rate (CFR) reported by Celentano et al. [6], equal to 6.3 per 1000 among infants younger than one year. We interpret this CFR as a proxy for the conditional probability *ρ*_cfr_ that an infected infant dies from pertussis before leaving the infectious state. This assumption, commonly adopted in compartmental epidemic models [43], gives *ρ*_cfr_ = 6.3 · 10^−3^. In the discrete–time setting, *ρ*_cfr_ is linked to the weekly survival probability *r*_3_ as follows. In each week, an infected infant dies with probability 1 − *r*_3_. If the individual survives, then he leaves the compartment *I*_1_ either by recovery or by ageing into the non–infant infectious class, with probability *γ*_1_ + *η*; otherwise, the individual remains in *I*_1_ with probability 1 −(*γ*_1_ + *η*). Therefore, the probability that death occurs in week *k* + 1, after *k* consecutive weeks spent in *I*_1_, is

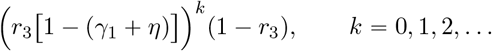

Summing over all possible weeks gives the geometric series

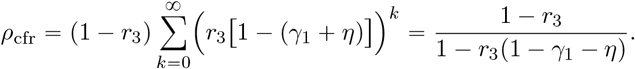

Solving for *r*_3_ yields

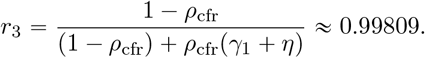

The values of 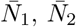, and *r*_1_ are then used to determine the recruitment rate Λ and the weekly survival probability of non–infants, *r*_2_, so that the demographic equilibrium matches the 2024 Italian population structure. At the demographic equilibrium, the age–specific population sizes can be expressed in terms of the model parameters as

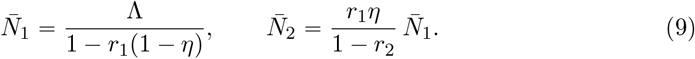

Thus, the weekly recruitment is

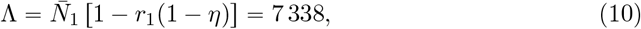

in line with the ISTAT estimates of approximately 1 040 births per day, that is, approximately 7280 per week [25]. Substituting into (9) yields

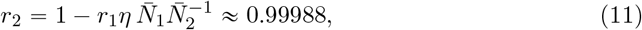

and the corresponding annual probability of death for non–infants is 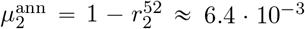. Finally, the time period spent in the first age group is 52 weeks. To derive the weekly probability of moving to the second age group, we use again the standard discrete–time conversion [42] and convert the ageing rate into the weekly probability of ageing:

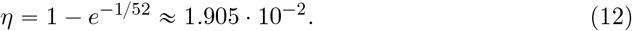

### 4.2 Vaccination–related parameters

Both the vaccination coverage and the vaccine effectiveness are parametrised from published sources. Poeta et al. [46] report that 62.6% of newborns are protected via maternal immunisation; accordingly, we set *p* = 0.626. According to the Istituto Superiore di Sanità (ISS), 94.7% of infants receive the vaccine annually [27]. We take 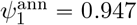 and convert it into a weekly probability,

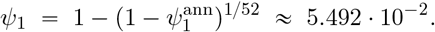

As for non–infant boosters, Lapi et al. [32] report an uptake of 16.5 booster doses per 10 000 individuals per year. In the model, we encode this quantity through the annual per–capita booster probability 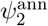, so that 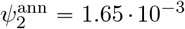 (i.e., 0.165% per year). For ease of interpretation, we sometimes express this booster uptake on the scale “per 10 000 individuals per year”, i.e., as the expected annual number of booster doses per 10 000 individuals. This is obtained by multiplying the per–capita probability by 10 000. The corresponding weekly probability is

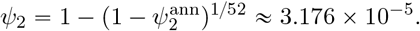

Imperfect protection is described by the vaccine failure probabilities *σ*_1_ (infants) and *σ*_2_ (non–infants). The infant compartment *V*_1_ comprises those protected by maternal immunisation and those who have received the second and third doses. In the absence of composition data, we use the arithmetic mean of the reported efficacies, i. e., 75.3% and 83.5% for the second and third DTaP doses, respectively [48], and 90% for maternal immunisation [3]. This yields an average efficacy of approximately 0.83, and thus *σ*_1_ = 1 − 0.83 = 0.17. Concerning the non–infant group, we use the estimated Tdap booster effectiveness, which is 92% [63], as a proxy for the vaccine-induced protection in the compartment *V*_2_. This yields an ineffectiveness factor of *σ*_2_ = 1 − 0.92 = 0.08.

The waning probability of vaccine–induced protection is informed by Schwartz et al. [51], who report a significant loss of protection within a decade, with estimates declining from 84% at 1–3 years since the last dose to 62% at 4–7 years and 41% after 8 years. Consistent with national recommendations for a decennial booster [29], we set the mean duration of vaccine–induced immunity to 10 years (520 weeks). This gives a weekly probability of losing vaccine–induced immunity

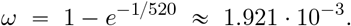

### 4.3 Epidemiological parameters

We assume the same mean infectious period in both age groups. According to the *Red Book 2025* [10], untreated infection lasts about 3 weeks. Thus, the corresponding weekly recovery probability is

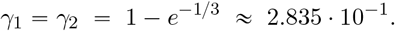

We further assume that infection–acquired immunity has the same mean duration as vaccine–induced immunity, namely 10 years (520 weeks), consistent with the 4–12 year range reported by Wendelboe et al. [66]. Hence, the weekly probability of losing infection–acquired immunity is

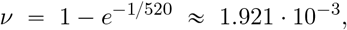

which matches the waning rate adopted for vaccine–induced immunity. As for the transmission rates, we assumed their values to obtain age–specific annual cumulative number of cases that are consistent with the estimates reported by the Istituto Superiore di Sanità and the World Health Organization [28, 67]. Hence, we set

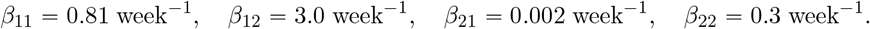

### 4.4 Initial conditions

We set the initial conditions in each age group so that 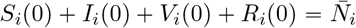, *i* = 1, 2. Since publicly available Italian pertussis data are reported as annual cumulative data, the initial data are assumed. We assume that the initial number of infected is *I*_1_(0) = 5 and *I*_2_(0) = 15. For vaccinated infants, the initial condition *V*_1_(0) includes those under 2 months protected by maternal immunisation, those aged 2–4 months who received maternal vaccination and also the first DTaP dose, and those who, by January 2024, had already received the second DTaP dose. We therefore approximate

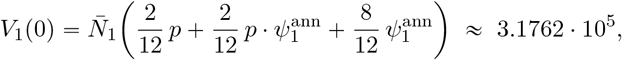

where *p* is the fraction of newborns protected via maternal vaccination and 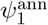 is the annual DTaP coverage in infants under 1 year of age. For the initial conditions of vaccinated non–infants, *V*_2_(0), we do not explicitly account for immunity due to recent non-infant booster doses, which is expected to be limited given the well–documented difficulties in achieving high booster uptake in adolescents and adults [16]. Moreover, as far as we know, detailed time series on booster uptake in adolescents and adults are not routinely available at the national level [39]. As a consequence, we cannot reconstruct, from data, the fraction of non–infants who received a booster in the years preceding *t* = 0, and we do not include this contribution when setting the initial condition *V*_2_(0). We therefore assume that *V*_2_(0) consists of children aged 1-10 years who completed the primary DTaP series during their first year of life and have not yet lost vaccine–induced immunity. To estimate this quantity, we extract from the demographic tables of ISTAT [26] the numbers of individuals aged *a* = 1, …, 10 years as of January 2024 and, for each cohort, multiply by the infant DTaP coverage recorded in its first year of life. We denote by *N*_*a*_ the size of the *a*-year-old cohort and by 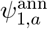 the corresponding infant coverage for the cohort’s birth year *y*_*a*_. The quantities *N*_*a*_ and 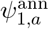^n^ are reported in Table 3 and are extracted from [26, 27]. In this way, we obtain:

**Table 3:** Cohort sizes, *N*_*a*_, and infant DTaP coverage, 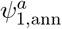, by birth year. The data about the resident population are available online on the ISTAT website [26], the temporal series of vaccination coverage is available online on the ISS website [27].

| Birth year | 2023 | 2022 | 2021 | 2020 | 2019 | 2018 | 2017 | 2016 | 2015 | 2014 |
| --- | --- | --- | --- | --- | --- | --- | --- | --- | --- | --- |
| $N_a$ | 397 193 | 407 572 | 414 581 | 431 500 | 450 440 | 469 364 | 483 663 | 495 818 | 510 387 | 517 826 |
| $\psi_{1,\text{ann}}^a$ | 94.76 % | 95.14 % | 94.00 % | 94.03 % | 94.99 % | 95.07 % | 94.62 % | 93.55 % | 93.33 % | 94.64 % |

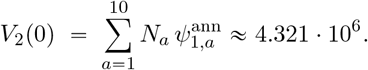

Regarding the initial condition for recovered individuals, we initialise at the onset of the epidemic wave by setting *R*_1_(0) = 0. As for non–infants, *R*_2_(0) represents individuals immune due to prior infection. Consistent with the assumption that infection–induced immunity lasts approximately ten years, *R*_2_(0) can be approximated by the number of pertussis cases recorded in the ten years preceding the initial time of the simulation, that is, January 2024. In practice, however, the ISS provides online cumulative incidence data only for 2014–2018 [28]. We therefore use the available series as a proxy for the last ten years and set *R*_2_(0) = 4064. Finally, the initial conditions for susceptibles are obtained by using the age–specific mass–balance constraint

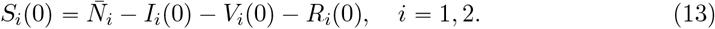

## 5 PerTexP outcomes

We use the MATLAB–based tool `PerTexP` to explore and compare different vaccination scenarios. The graphical user interface of `PerTexP` is described in detail in Section 5.5, and its code is available via a public GitHub repository (https://github.com/emanuelapenitente/PerTexP). Simulations were performed using a reduced system with state variables *S*_1_(*t*), *V*_1_(*t*), *I*_1_(*t*), *S*_2_(*t*), *V*_2_(*t*), *I*_2_(*t*), that is equations (3a)-(3c) and (3e)-(3g), while the size of the recovered compartment was computed a posteriori at each time step by using the age-specific mass balance constraint

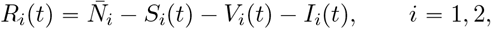

where 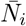 denotes the 2024 Italian population size in the *i*–th age group, assumed constant over the considered time horizon.

Before presenting the results of these scenarios, we define the following quantities used to evaluate the impact of each intervention:

- The *annual cumulative incidence* in the *i*–th age group during year *y*, denoted by CI_*i*_(*y*). This quantity represents the cumulative number of pertussis cases occurring during year *y* in age group *i*. It is computed by summing, over the 52 weeks of year *y*, the weekly number of new infections in age group *i*. In the model, new infections occurring between weeks *t* and *t* + 1 are given by the infection-related inflow into the infected compartment, i.e., *r*_*i*_*G*_*i*_(*t*)[*S*_*i*_(*t*) + *σ*_*i*_*V*_*i*_(*t*)]. Therefore,

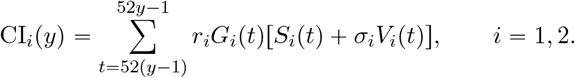
- The *annual cumulative number of vaccinations* administered in the *i*–th age group during year *y*, denoted by CV_*i*_(*y*). This quantity is computed by summing, over the 52 weeks of year *y*, the weekly number of newly vaccinated individuals in age group *i*. In the model, weekly vaccinations correspond to the vaccination-related inflow into the vaccinated compartment *V*_*i*_ between weeks *t* and *t* + 1. For infants (*i* = 1), this includes both newborns protected at birth through maternal vaccination, represented by the term *p*Λ, and vaccinations administered to susceptible infants, represented by the term *ψ*_1_*S*_1_(*t*). For non–infants (*i* = 2), vaccinations are given by *ψ*_2_*S*_2_(*t*). Therefore,

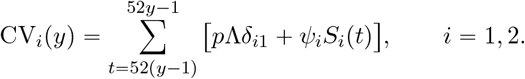

where *δ*_*i*1_ denotes the Kronecker delta, i.e., *δ*_*i*1_ = 1 if *i* = 1 and *δ*_*i*1_ = 0 otherwise.

### 5.1 Exploring the 2024 Italian pertussis outbreak

We first provide a five–year projection of pertussis transmission in Italy based on the 2024 demographic and clinical parameters reported in Table 2. Figure 2 shows the annual cumulative incidence for infants (blue bars) and non–infants (orange bars), while Figure 3 reports the corresponding annual cumulative vaccinations, distinguishing immunisation of infants (Figure 3a) and non–infant administration of booster doses (Figure 3b). Infant vaccinations are reported as a percentage of the total infant population, whereas booster doses are reported per 10 000 individuals.

**Figure 2:**
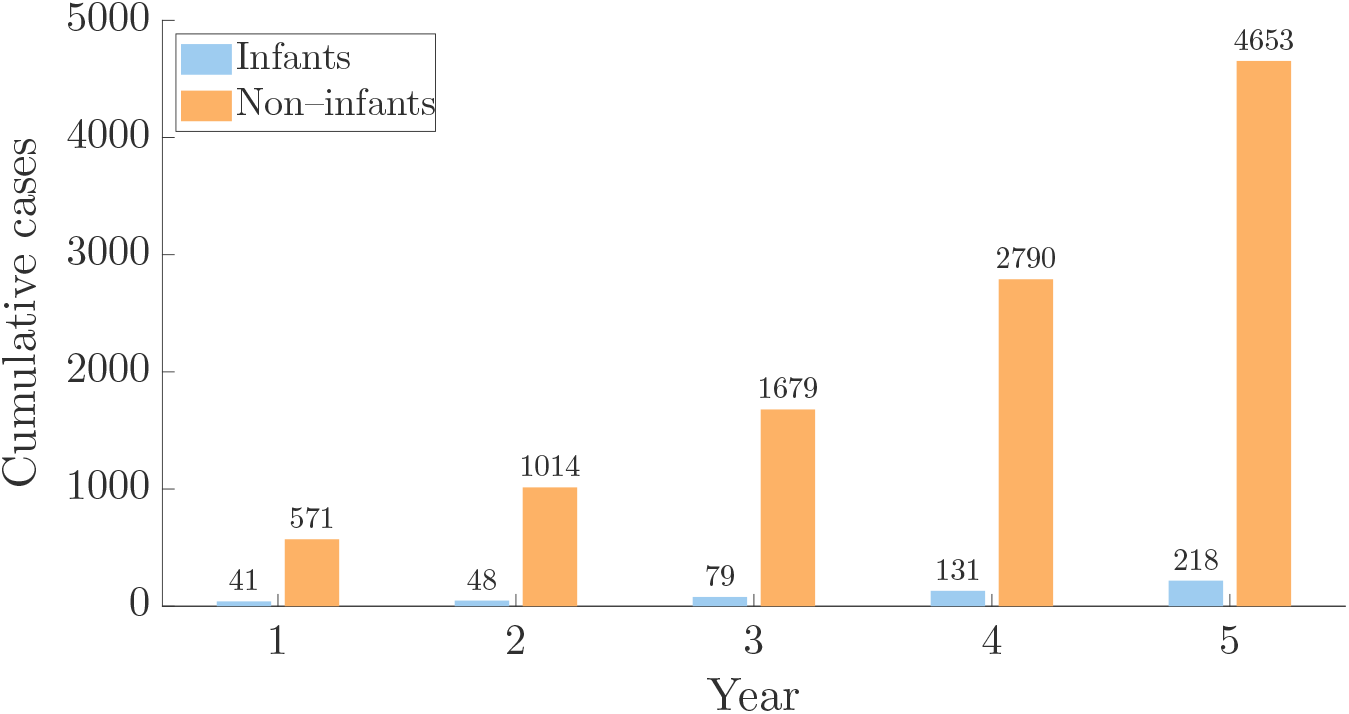
Five-year projection of pertussis transmission in Italy obtained with `PerTexP`, under the 2024 demographic and epidemiological parameters in Table 2. Bars report the annual cumulative number of new pertussis cases for infants (blue bars) and non–infants (orange bars).

**Figure 3:**
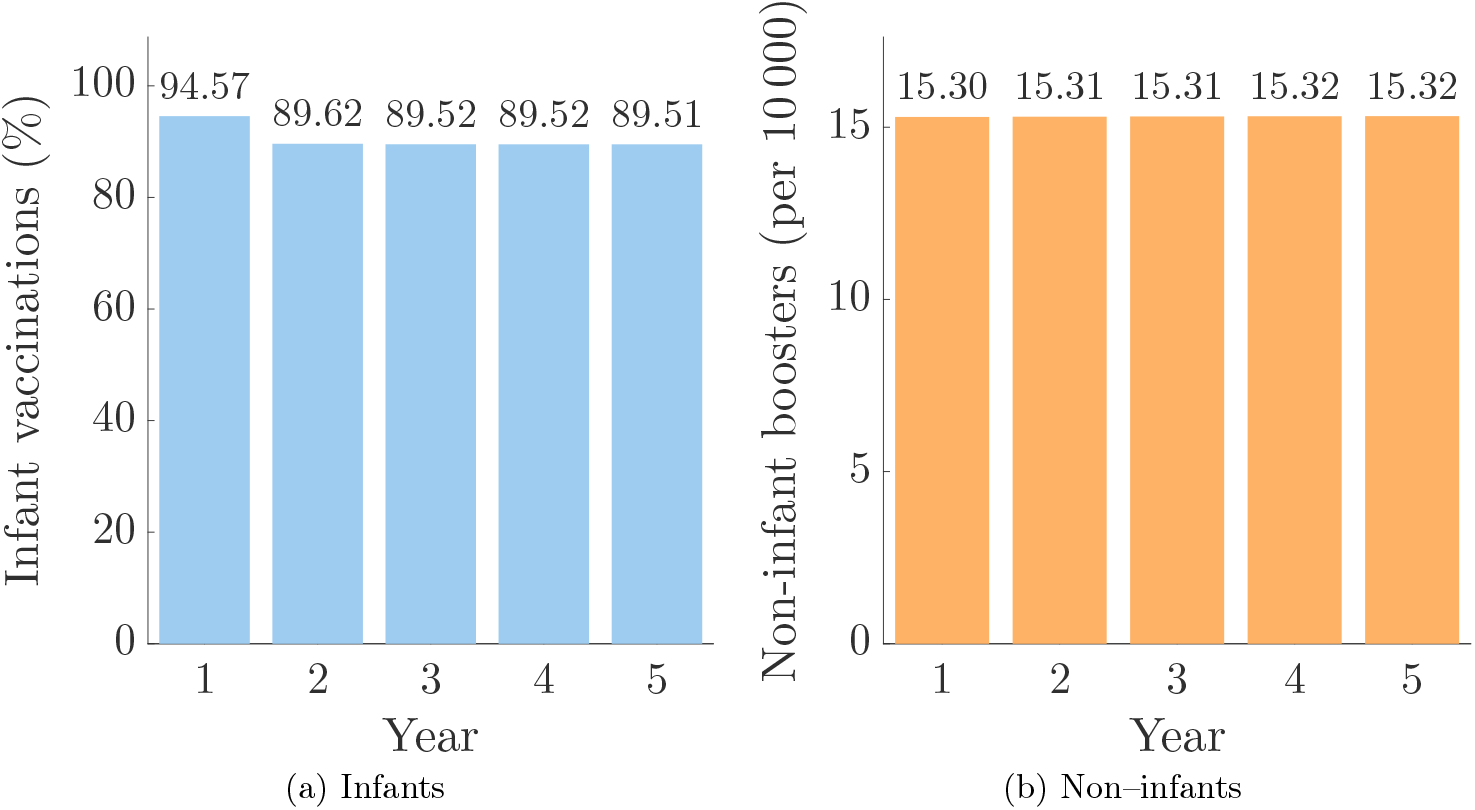
Annual cumulative numbers of infant vaccinations and non–infant booster doses simulated by `PerTexP` over five years. Panel (a) shows maternal and infant primary vaccinations, while panel (b) shows non–infant boosters.

The annual pertussis incidence reported in Figure 2 presents an increasing trend over the five-year horizon, rising from 612 total cases in the first year to 4871 in the fifth year. Infant cases range from 41 in the first year to 218 in the fifth year. A direct comparison with the 2024 Italian outbreak is not straightforward since the published data include only hospitalised infants, collected in a limited number of hospitals and over a restricted period [46]. However, these data still provide a useful reference for severe disease and suggest that our projections are in a plausible range. Overall, our estimates should be seen as conservative, since the true infant incidence also includes subclinical and non-hospitalised infections. Similarly, the projected total number of cases appears to be broadly consistent with the World Health Organization (WHO) data reported for Italy [67].

Note that infant infections account for a small fraction of the total cases (approximately 5%). This is consistent with the fact that infants under 1 year of age represent only a small fraction of the total Italian population (approximately 0.65%). However, infant infections remain epidemiologically relevant because disease severity is significantly higher in this age group, with an increased risk of complications such as pneumonia, neurological impairment and even death.

In these simulations, the pertussis outbreak appears to have only a negligible effect on the vaccination campaign, which shows a stable trend throughout the simulation period (see Figure 3). This is consistent with the fact that the number of cases is relatively small compared to the total population, so the depletion of susceptibles (and hence annual cumulative vaccinations) remains minimal.

### 5.2 Impact of vaccination coverages on pertussis transmission

We investigate how the vaccination–related parameters, namely the annual infant vaccination coverage 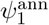, the annual booster coverage 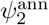 and the maternal immunization coverage *p*, influence pertussis transmission. More precisely, we assess how varying these parameters affects the control reproduction number, whose unit threshold is the dividing line between the elimination and the onset of the epidemic. The results are shown in Figure 4, which displays contour plots of ℛ_*c*_ versus 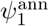 and *p* (panel 4a), and versus 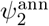 and *p* (panel 4b). Each isoline denotes points with the same ℛ_*c*_ value, while the dotted white lines indicate the values corresponding to the 2024 Italian scenario, in which ℛ_*c*_ = 1.037. The black curve in panel 4b is the isocline where ℛ_*c*_ = 1. As expected, the control number decreases monotonically as each vaccination coverage increases, attaining its minimum when both couples of strategies are implemented at the maximum values in the explored range. However, the progressively wider distance between the contour lines in the upper-right region of panel 4a suggests that the reduction in ℛ_*c*_ saturates at high maternal and infant immunisation coverage. The 2024 Italian scenario lies in this region, suggesting a negligible reduction in ℛ_*c*_ from further increases in infant or maternal coverage. Panel 4b suggests that, within the explored range 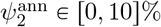, the sensitivity of ℛ_*c*_ to the booster coverage is not uniform. At very low booster uptake, changes in 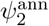 have a significant impact on _*c*_, whereas the contour lines become progressively flatter as 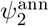 increases (e.g., if 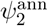 is approximately above 5%), indicating a stronger sensitivity to the maternal coverage *p* in that region. However, increasing *p* alone is not sufficient to achieve pertussis elimination: attaining ℛ_*c*_ < 1 requires increasing both maternal immunisation and booster coverage.

**Figure 4:**
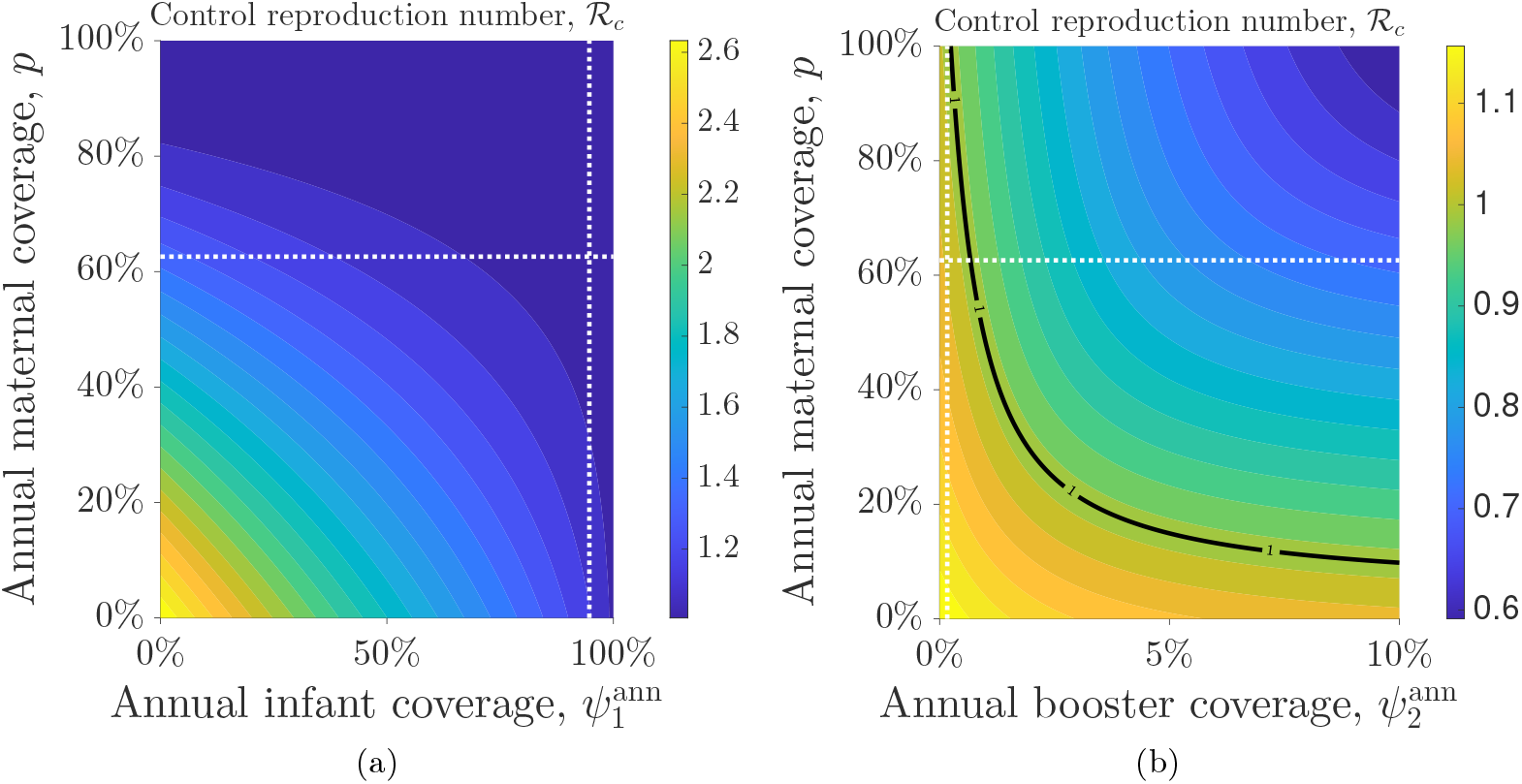
Contour plots of the control reproduction number versus the vaccination–related parameters. Panel (a) shows ℛ_*c*_ versus 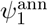 and *p*. Panel (b) shows ℛ_*c*_ versus 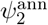 and *p*. The intersection between the dotted white lines indicates the values corresponding to the 2024 Italian scenario, namely, *p* = 62.6%, 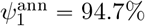, and 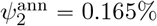 (i.e., 16.5 per 10 000 per year). All other parameters are fixed as in Table 2.

### 5.3 Impact of increasing booster coverage versus maternal immunisation coverage on pertussis cumulative cases

We compare three vaccination scenarios to assess their effectiveness in reducing the cumulative cases of pertussis:

i. the 2024 Italian scenario, considered as the baseline scenario, with parameter values fixed as in Table 2;
ii. the scenario in which the annual booster coverage 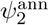 is increased by 10% compared to its 2024 baseline value, yielding 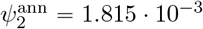 (i.e., 18.15 booster doses per 10 000 individuals per year);
iii. the scenario in which maternal immunisation coverage *p* is increased by 10% compared to its baseline value, yielding *p* = 0.689.

In the last two scenarios, all the remaining parameters are held at their baseline values reported in Table 2. For each scenario, we evaluate the relative changes in the 5–year cumulative incidence among infants, CI_1_, among non–infants, CI_2_, and across both age groups, CI_tot_, with respect to the baseline scenario. To this end, we define the index

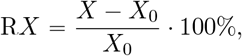

which quantifies the percentage *relative* change of *X* ∈ tCI_1_, CI_2_, CI_tot_u with respect to the corresponding quantity *X*_0_ predicted by `PerTexP` in the Italian 2024 baseline scenario, with 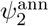 and *p* set as in Table 2.

Table 4 illustrates the results, and Figure 5 shows the annual cumulative cases predicted by `PerTexP` for the three scenarios. Panel 5a illustrates the projected annual cumulative incidence for infants, whereas panel 5b refers to non–infants. In both panels, the green bars represent scenario (*ii*), while the blue bars refer to scenario (*ii*). The grey bars represent the Italian context in 2024, denoted as scenario (*i*). Table 4 and Figure 5 show that both the increase in booster coverage and the increase in maternal coverage reduce the annual cumulative incidence of both infants and non–infants compared to the Italian scenario for 2024. However, increasing only the booster coverage results in a reduction of approximately 1.2% in cumulative cases over five years in both age groups. By contrast, increasing the maternal coverage *p* has a significantly larger effect: infants experience 37.2% fewer cases than in the baseline scenario, and non–infants also indirectly benefit from this strategy, with a projected 29.4% reduction in cumulative cases over the same time period.

**Table 4:** Five-year cumulative incidence among infants, CI_1_, among non–infants, CI_2_, and total, CI_tot_, under three combinations of vaccination–related parameters *p* and 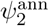, together with the corresponding relative change with respect to the 2024 Italian scenario (first row). All other parameter values are reported in Table 2.

| Scenario | | $CI_1$ | $RCI_1$ | $CI_2$ | $RCI_2$ | $CI_{\text{tot}}$ | $RCI_{\text{tot}}$ |
| --- | --- | --- | --- | --- | --- | --- | --- |
| Italy, 2024 (baseline) | (i) | 517 | 0 | 10 708 | 0 | 11 224 | 0 |
| +10% booster coverage | (ii) | 511 | −1.10 | 10 249 | −1.22 | 11 088 | −1.21 |
| +10% maternal coverage | (iii) | 321 | −37.85 | 7 524 | −29.7 | 7 845 | −30.10 |

**Figure 5:**
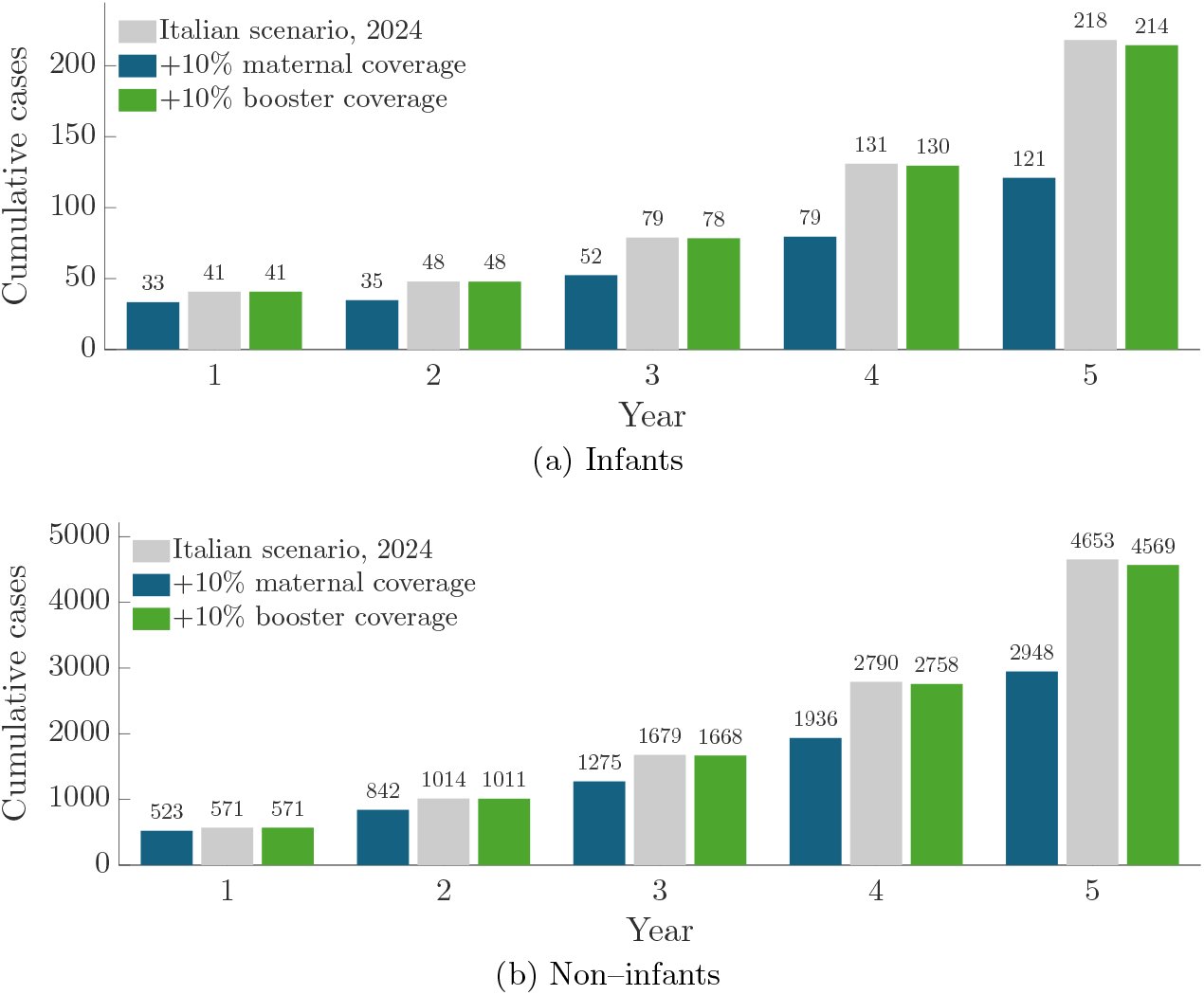
Comparison of two intervention scenarios over a five-year horizon. Bars report the annual cumulative number of new pertussis cases in (a) infants and (b) non–infants. The blue bars correspond to a 10% increase in the maternal immunisation coverage *p* compared to its 2024 Italian value, while the green bars correspond to a 10% increase in the annual non–infant booster coverage 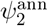. All the other parameters are fixed to their baseline values reported in Table 2.

### 5.4 Effects of random perturbations on PerTexP outcomes

The model underlying `PerTexP` is governed by a system of deterministic equations: for a fixed set of parameters and initial conditions, the system evolves in a fully prescribed way, so that repeated `PerTexP` runs from the same initial conditions yield the same outputs. In this sense, `PerTexP` describes the average course of the outbreak and is useful for investigating the underlying epidemic dynamics and comparing different policy scenarios. We complement the deterministic analysis by investigating the role played by randomness, particularly relevant whenever the number of infectious individuals is relatively small [31]. We introduce randomness into the model by adding small random fluctuations in the state variables at the end of each year, in order to assess the robustness of `PerTexP` to small year-to-year variability in the system state. Such random perturbations aim to represent the effect of unobserved factors that are not explicitly captured by the deterministic model structure. At the end of each year (i.e., at *t* = 52*y* for *y* = 1, …, 4), the state variables **x**(*t*) = *S*_1_(*t*), *V*_1_(*t*), *I*_1_(*t*), *S*_2_(*t*), *V*_2_(*t*), *I*_2_(*t*) of the reduced system (3a)–(3c) and (3e)–(3g) are randomly perturbed through multiplication with scaling parameters *ε*_1_, …, *ε*_6_, where the scaling parameters *ε*_*j*_ are independent and generated from a Gaussian distribution, 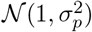, with *σ*_*p*_ = 5% controlling the magnitude of the fluctuations. At the end of each year, each state variable is then perturbed as

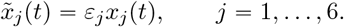

The perturbed state variables vector 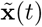 is then used as initial condition for the simulation over the following year, and the procedure is repeated again for every year.

Since the state variables 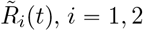, are computed implicitly as 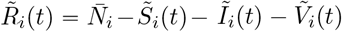, we ensured their nonnegativity by applying an age–specific renormalisation whenever the sum of the perturbed susceptible, vaccinated and infected individuals exceeds the corresponding population size. Specifically, denoting

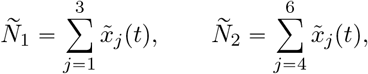

we set

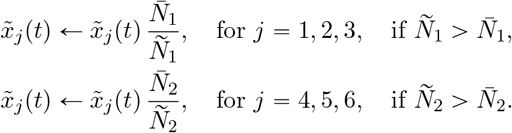

Figure 6 illustrates the effect of these random perturbations on the annual cumulative pertussis incidence for a single realisation of the scaling parameters *ε*_1_, …, *ε*_6_. Introducing these perturbations can yield a non monotone profile in the cumulative cases, particularly in the non–infant group, while preserving the qualitative behaviour of the deterministic projection. In this example, annual cumulative incidence remains within the range of values projected under the baseline scenario and does not exceed the baseline projection.

**Figure 6:**
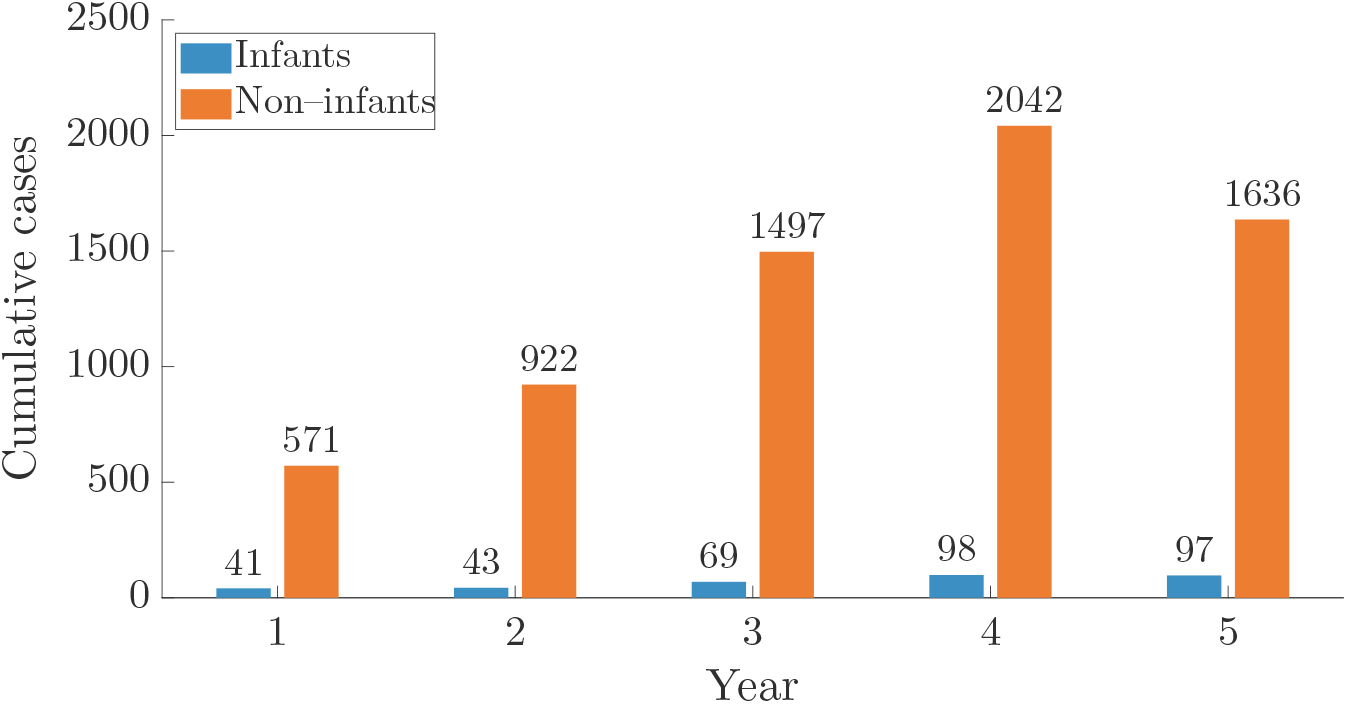
Five-year projection of annual cumulative pertussis incidence in Italy obtained with `PerTexP`. Model parameters and baseline initial conditions at the beginning of the first year are reported in Table 2. At the end of each year, the state variables of the reduced system (3a)–(3c) and (3e)–(3g) were randomly perturbed through multiplicative noise parameters, which are independent and generated from a Gaussian distribution 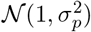, with *σ*_*p*_ = 0.05. The perturbed state variables are used as initial conditions for the following year. Blue bars show the annual cumulative number of new pertussis cases in infants, while orange bars refer to non–infants.

**Figure 7:**
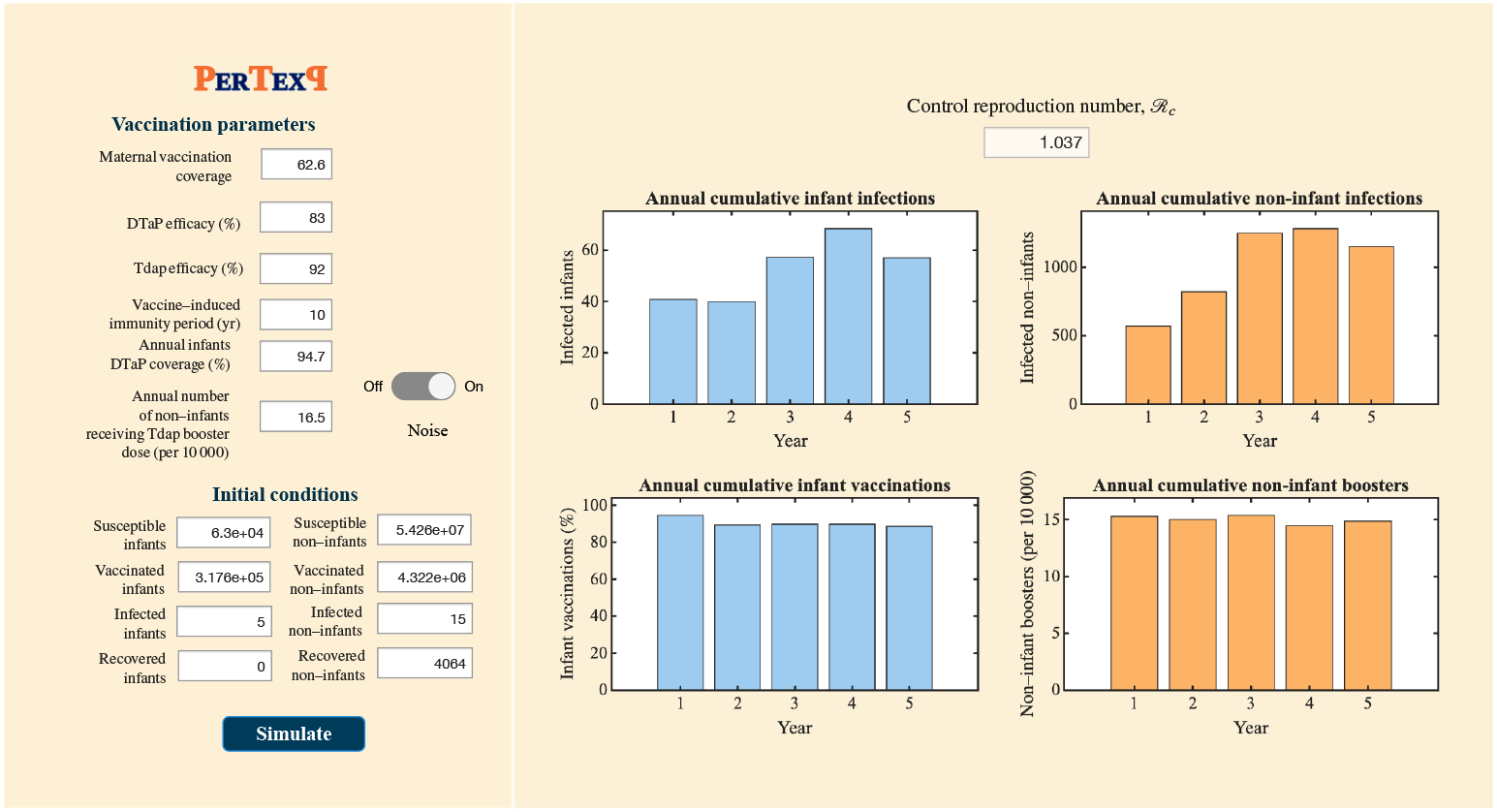
`PerTexP` graphical user interface implemented in MATLAB App Designer. The input panel (left) allows specification of vaccination parameters and initial conditions for infants and non–infants; the output panel (right) reports the value of the control reproduction number, together with the age–specific annual cumulative pertussis infections and vaccinations over a five–year horizon.

### 5.5 PerTexP graphical user interface

`PerTexP` is implemented as a MATLAB app with a simple Graphical User Interface (GUI), whose layout separates user inputs (left panel) from model outputs (right panel), enabling rapid scenario exploration. The app was developed using MATLAB *App Designer*, the integrated environment for building interactive applications, available in MATLAB since release R2016a. The source code, including the GUI implementation, is publicly available on GitHub (https://github.com/emanuelapenitente/PerTexP). `PerTexP` is novice–friendly, and experience in programming with MATLAB is not required. The input panel is organised into two sections: the *Vaccination parameters* section allows users to set the maternal immunisation coverage *p*, both the infant DTaP and non–infant Tdap coverages and efficacies, and the duration of the vaccine–induced protection, whereas the *Initial conditions* section allows users to specify the starting values of the epidemiological compartments in each age group (infants and non–infants). An optional *Noise* switch enables the annual perturbation procedure described in Section 5.4, allowing users to assess how small year–to–year fluctuations in the system state affect projected outcomes. Once inputs are set, clicking *Simulate* runs the model over the five–year horizon and updates the output panel. The latter displays the value of the control reproduction number ℛ_*c*_, the age–specific annual cumulative pertussis incidence and the annual cumulative vaccinations over time, providing an immediate visual exploration of different vaccination scenarios.

## 6 Concluding remarks

Our study concerns the formulation and implementation of `PerTexP` (Pertussis Time Exploration), a MATLAB–based tool designed to explore pertussis transmission dynamics under maternal and infant vaccination strategies. `PerTexP` is built upon a discrete–time compartmental model with two age groups, in which transmission within each group follows a SIR–type structure and vaccine–induced immunity is modelled through an additional vaccinated compartment in each age class. The aim of `PerTexP` is to allow the exploration and comparison of alternative vaccination strategies. Through its graphical user interface, users can specify vaccination–related parameters and initial conditions, and receive as output age–specific annual cumulative incidences over a five–year time horizon. We focus on the 2024 Italian pertussis outbreak as an illustrative case study, parametrising `PerTexP` to reproduce annual cumulative incidences consistent with those reported during the epidemic. Some examples of the potential of `PerTexP` are as follows:

- Projecting age–specific pertussis annual cumulative incidence over a multi–year horizon, helping users to better understand the current trend of the epidemic. For example, when applied to the 2024 Italian outbreak, `PerTexP` projects an increasing pertussis annual cumulative incidence over a five–year horizon, with the total cases rising from 606 in the first year to 4682 in the fifth year.
- Assessing the sensitivity of key epidemiological indicators, such as the control reproduction number ℛ_*c*_, to vaccination–related parameters, and identifying intervention combinations required for disease elimination. In the Italian case study, `PerTexP` suggests that ℛ_*c*_ is significantly more sensitive to variations in maternal immunisation coverage than to booster coverage; however, maternal coverage alone is not sufficient to reduce ℛ_*c*_ below one, and pertussis elimination requires a combined increase in both maternal immunisation and booster uptake.
- Quantifying and comparing the impact of different vaccination strategies on cumulative incidence. In the 2024 Italian case study, `PerTexP` indicates that increasing maternal immunisation yields significantly larger reductions in cumulative incidence than increasing decennial booster uptake. Boosters mainly reduce incidence among non–infants and contribute indirectly to the epidemic control by lowering the control reproduction number, with a comparatively modest impact on infant incidence. Thus, booster uptake appears to be a supporting strategy to reduce infant pertussis burden.

To further support the reliability of `PerTexP`, we note that the comparative insights on maternal immunisation and booster uptake obtained in the Italian scenario are consistent with those obtained with other modelling approaches and from other case studies [5, 15, 21, 44, 56]. Moreover, they align with recent public health recommendations; in fact, the European Centre for Disease Prevention and Control highlights maternal vaccination as an effective strategy to protect newborns, while noting that decennial boosters mainly benefit older age groups and should be regarded as complementary for infant protection [14].

We acknowledge that `PerTexP` presents some limitations, which nevertheless leave the possibility of future developments. In particular, a first limitation concerns the range of inputs that can be modified through `PerTexP`. In the current release, users can change vaccination–related parameters and initial conditions, whereas demographic parameters are kept fixed and cannot be edited via the interface. This choice is not due to a lack of generality of the underlying model, as `PerTexP` can be applied to any geographical region, but rather stems from the way in which transmission rates are selected. In the absence of weekly time series of infected prevalence and/or incidence to calibrate the model, the transmission rates are not inferred by fitting the model to the temporal dynamics of the outbreak. They are assumed to reproduce the age–specific annual cumulative incidence values that are consistent with the 2024 Italian scenario. Allowing demographic parameters to vary without a corresponding recalibration of the transmission rates could therefore generate outcomes that are not epidemiologically realistic. For this reason, demographic inputs are not made tunable in the current version of `PerTexP`. While acknowledging that transmission parameters are not inferred from time–series data, the adopted values yield epidemiologically plausible `PerTexP` outcomes in terms of overall burden and provide an illustrative example of how the tool can be parametrised for a specific case study.

A second limitation involves neglecting the element “human behaviour”: in some cases, vaccine uptake and contact patterns may vary in response to perceived epidemic risk and fear of vaccine–side effects. Incorporating data–driven calibration and behavioural changes would therefore be valuable extensions to improve the reliability of `PerTexP`.

## Acknowledgements

This work has been carried out under the auspices of the Italian National Group for Mathematical Physics (GNFM) of the National Institute for Advanced Mathematics (INdAM). The research was supported by EU funding within the NextGenerationEU–MUR PNRR Extended Partnership initiative on Emerging Infectious Diseases (Project no. PE00000007, INF-ACT).

## Data availability statement

Data sharing is not applicable to this article as no datasets were generated or analysed during the current study.

## Compliance with ethical standards

### Conflict of interest

The authors state that there is no conflict of interest.

## Notes

### Competing Interest Statement

The authors have declared no competing interest.

