## Supplementary material for "PerTexP: scenario-based exploration of pertussis dynamics under maternal and infant vaccination"

### S.1 The contagion probability

The choice of the contagion probability function $G_i(\cdot)$ has a significant impact on both the formulation and the model results [4]. Several functional forms have been proposed in the literature on discrete–time models, and some authors regard it as a modelling paradigm [2]. One possible approach, known as *frequency-dependent*, assumes that $G_i(\cdot)$ depends on the proportion of infectious individuals in the total population. An alternative approach is the *density-dependent* formulation, in which $G_i(\cdot)$ depends on the absolute number of infectious individuals. The term “density-dependent” reflects the fact that the probability of contagion is determined by the number of infectious individuals per unit of space, that is, by the density of infectious individuals. As explained in Section 2.3 of the main text, we model transmission using a frequency-dependent formulation, that is, the per-susceptible risk of infection due to infectious individuals of the $j$–th age group scales with the prevalence $I_j/N_j$. Indeed, frequency-dependent transmission is considered more appropriate for modelling pathogens that spread through a heterogeneous contact structure [9]. It is also consistent with continuous age-structured pertussis models [8] and with discrete-time age-structured models of other airborne infections where infants play a key role [12]. Since the transmission occurs through contact between susceptible and infectious individuals across all age groups, we assume that $G_i = G_i(\lambda_i)$, where

$$\lambda_i = \sum_{j=1}^{2} \beta_{ij} \frac{I_j(t)}{N_j(t)}, \quad i = 1, 2, \tag{1}$$

where the age-specific transmission rate $\beta_{ij} = c_{ij}\pi_{ij}$ is defined as the product of the average number of contacts per time step $c_{ij}$ between individuals in age groups $i$ and $j$, and the probability $\pi_{ij}$ of successful transmission given contact with an infectious individual from group $j$. The contact structure is represented by a contact matrix $\mathbf{C} = (c_{ij})_{i,j=1}^2$, where each element $c_{ij}$ denotes the average number of contacts per time step that an individual in age group $i$ has with individuals in age group $j$.

As for the functional form of the contagion probability $G_i(\lambda_i)$, we require it to satisfy the following properties [3]: $(i)$ $G_i\colon [0, +\infty) \to [0, 1]$; $(ii)$ $G_i(0) = 0$; $(iii)$ $G_i'(\lambda_i) > 0$. In our formulation, infections are modelled as Poisson processes, namely, the so–called "*infectious events*" (i.e., the infectious contacts that a susceptible experiences over a given time interval) occur randomly and independently, with an average number $\lambda_i$ of events per individual per time interval [6]. This implies that the random variable $X_i$ measuring the number of "infectious events" for a susceptible $S_i$ in a given time interval follows a Poisson distribution with mean $\lambda_i$, with a discrete probability distribution given by

$$p_{X_i}(k) = \frac{{\lambda_i}^k e^{-\lambda_i}}{k!}, \quad k \in \mathbb{N}_0.$$

The *escape probability* $\Phi_i(\lambda_i)$, i.e., the probability that an individual of the age class $i$ remains susceptible at the next time step, corresponds to the probability of having zero infectious events in the time interval $[t, t+1)$, therefore

$$\Phi_i(\lambda_i) = p_{X_i}(0) = e^{-\lambda_i}.$$

Thus, the *contagion function* is given by:

$$G_i(\lambda_i) = 1 - e^{-\lambda_i}.$$

Finally, the weekly average number $\lambda_i$ of infectious contacts per susceptible in the $i$–th age group is given in equation (1).

### S.2 Basic properties of the model

Model (3) – (4) can be rewritten in the vector form as

$$\mathbf{x}(t+1) = \mathbf{F}(\mathbf{x}(t)), \qquad \mathbf{x}(0) = \mathbf{x}_0,$$

where $\mathbf{x} = \big(S_1, V_1, I_1, R_1, S_2, V_2, I_2, R_2\big)$ denotes the state variable vector, $\mathbf{F}\colon \mathbb{R}^8 \to \mathbb{R}^8$ is the autonomous vector field and $\mathbf{x}_0$ is the vector of the initial conditions. We first establish the positivity of solutions under biologically meaningful parameter assumptions.

**Lemma 1.** Assume that the following inequality holds for all $t \in \mathbb{N}$:

$$\begin{aligned} 0 <\gamma_1 + \eta < 1, \quad 0 < G_1(t) + \psi_1 + \eta < 1, \quad 0 < \sigma_1 G_1(t) + \eta < 1, \\ 0 < G_2(t) + \psi_2 < 1, \quad 0 < \sigma_2 G_2(t) + \omega < 1. \end{aligned} \tag{2}$$

If the initial condition $\mathbf{x}_0 = \mathbf{x}(0)$ of model (3) has all strictly positive components, then the solution $\mathbf{x}(t)$ remains strictly positive for all $t \in \mathbb{N}$.

*Proof.* We proceed by induction on $t$. By hypothesis, the initial condition satisfies $\mathbf{x}(0) > 0$, and we prove that this implies $\mathbf{x}(t) > 0$ for all $t \in \mathbb{N}$. For $t = 1$, we have

$$S_1(1) = (1-p)\Lambda + r_1 \left[1 - G_1(0) - \psi_1 - \eta\right] S_1(0).$$

Since $\Lambda > 0$, $S_1(0) > 0$, $r_1 > 0$, and $p < 1$, it follows that $S_1(1) > 0$, under the parameter assumptions (2). Similarly, we obtain

$$V_1(1) = p\Lambda + r_1 \left[1 - \sigma_1 G_1(0) - \eta\right] V_1(0) + r_1 \psi_1 S_1(0) > 0,$$

$$\begin{aligned}
I_1(1) &= r_1 G_1(0)\left[S_1(0) + \sigma_1 V_1(0)\right] + r_3(1-\gamma_1-\eta) I_1(0) > 0, \\
R_1(1) &= r_3\gamma_1 I_1(0) + r_1(1-\eta)R_1(0) > 0, \\
S_2(1) &= r_1\eta S_1(0) + r_2\nu R_2(0) + r_2\left[1 - G_2(0) - \psi_2\right] S_2(0) + r_2\omega V_2(0) > 0, \\
V_2(1) &= r_1\eta V_1(0) + r_2\psi_2 S_2(0) + r_2\left[1 - \sigma_2 G_2(0) - \omega\right] V_2(0) > 0, \\
I_2(1) &= r_3\eta I_1(0) + r_2 G_2(0)\left[S_2(0) + \sigma_2 V_2(0)\right] + r_2(1-\gamma_2) I_2(0) > 0, \\
R_2(1) &= r_1\eta R_1(0) + r_2\gamma_2 I_2(0) + r_2(1-\nu)R_2(0) > 0.
\end{aligned}$$

Hence, $\mathbf{x}(1) > 0$. Assume now that $\mathbf{x}(t) > 0$ for some $t \in \mathbb{N}$. Then, from the system equations and under the same assumptions, we have

$$S_1(t+1) = (1-p)\Lambda + r_1\left[1 - G_1(t) - \psi_1 - \eta\right] S_1(t) > 0,$$

and all other components of $\mathbf{x}(t+1)$ are strictly positive by analogous reasoning. Therefore, $\mathbf{x}(t+1) > 0$, and the result follows by induction. □

**Proposition 1.** The region

$$\Omega = \left\{ \mathbf{x} \in \mathbb{R}^8_+ \,:\, \sum_{k=1}^{8} x_k \leqslant \frac{\Lambda}{\min\{\mu_1, \mu_2, \mu_3\}} \right\},$$

where $\mathbb{R}^8_+$ denotes the non–negative cone of $\mathbb{R}^8$, is positively invariant for model (3).

*Proof.* We want to prove that, if the initial condition $\mathbf{x}_0 = \mathbf{x}(0) \in \Omega$, then the solution $\mathbf{x}(t)$ remains in $\Omega$ for all $t \in \mathbb{N}$. From Lemma 1, it follows that any solution $\mathbf{x}(t)$ of model (3) satisfies $\mathbf{x}(t) \in \mathbb{R}^8_+$ for all $t$. Observe that, since $r_i = 1 - \mu_i$ for $i = 1, 2, 3$, we have

$$\min\{\mu_1, \mu_2, \mu_3\} = 1 - \max\{r_1, r_2, r_3\}.$$

We also recall that the total population is given by $N(t) = \sum_{i=1}^{8} x_i(t)$. Thus, our aim is to prove that, from

$$N(0) \leqslant \frac{\Lambda}{1 - \max\{r_1, r_2, r_3\}},$$

it follows that

$$N(t) \leqslant \frac{\Lambda}{1 - \max\{r_1, r_2, r_3\}} \quad \text{for all } t \in \mathbb{N}.$$

We proceed by induction on $t$. For $t = 1$, we have

$$N(1) = \Lambda + r_1 S_1(0) + r_1 V_1(0) + r_3 I_1(0) + r_1 R_1(0) + r_2 S_2(0) + r_2 V_2(0) + r_2 I_2(0) + r_2 R_2(0).$$

If we set $r := \max\{r_1, r_2, r_3\}$, then

$$N(1) \leqslant \Lambda + r\, N(0).$$

By hypothesis, $N(0) \leqslant \Lambda/(1-r)$, and therefore

$$N(1) \leqslant \Lambda + \frac{r\Lambda}{1-r} = \frac{\Lambda}{1-r}.$$

Assume now that $N(t) \leqslant \Lambda/(1-r)$ for some $t \in \mathbb{N}$. Then the same computation yields

$$N(t+1) \leqslant \Lambda + r\, N(t) \leqslant \Lambda + \frac{r\Lambda}{1-r} = \frac{\Lambda}{1-r}.$$

Therefore, $N(t) \in \Omega$ for all $t \in \mathbb{N}$, and we conclude that the region $\Omega$ is positively invariant. □

### S.3 Details on the computation of the reproduction numbers

As mentioned in Section 3 of the main text, we compute the expressions of the basic reproduction number $\mathcal{R}_0$ and the control reproduction number $\mathcal{R}_c$ by using the *next-generation matrix* approach. The underlying approach was originally introduced in 1990 by Diekmann et al. [5] for continuous–time models of heterogeneous populations, where $\mathcal{R}_0$ is defined as the spectral radius (that is, the dominant eigenvalue) of a positive linear operator, the *next-generation* operator. In 2002, van den Driessche and Watmough [13] provided a procedure to build the corresponding *next-generation matrix* (NGM) in the case of compartmental models governed by ordinary differential equations. For discrete–time models, this procedure was introduced by Allen and van den Driessche in 2008 [1]. According to the NGM approach, the discrete–time system (3) can be arranged as

$$\begin{cases} \mathbf{x}_1(t+1) \\ \mathbf{x}_2(t+1) \end{cases} = \begin{cases} \mathcal{F}(\mathbf{x}(t)) + \mathcal{T}(\mathbf{x}(t)) \\ \mathcal{H}(\mathbf{x}(t)) \end{cases},$$

where $\mathbf{x}_1 \in \mathbb{R}^{n_1}$ is the vector containing the state variables related to the infected compartments, i.e. $\mathbf{x}_1 = (I_1, I_2)$, and $\mathbf{x}_2 \in \mathbb{R}^{n_2}$ is the vector of the state variables related to the uninfected ones, that is, $\mathbf{x}_2 = (S_1, V_1, R_1, S_2, V_2, R_2)$. The vector $\mathcal{F}$ collects the terms that generate *new infections* and $\mathcal{T}$ is the vector collecting all other transitions (e.g., ageing, recovery or pertussis-related deaths). The Jacobian matrices of $\mathcal{F}$ and $\mathcal{T}$, evaluated at the disease–free equilibrium $E_0 = (0, \bar{\mathbf{x}}_2)$, are denoted as the *fertility* matrix $\mathbf{F}$ and *transition* matrix $\mathbf{T}$, respectively:

$$\mathbf{F} = \left[\frac{\partial \mathcal{F}_i}{\partial \mathbf{x}_{1,j}}(E_0)\right]_{i,j=1}^{n_1}, \qquad \mathbf{T} = \left[\frac{\partial \mathcal{T}_i}{\partial \mathbf{x}_{1,j}}(E_0)\right]_{i,j=1}^{n_1}.$$

Note that $\mathbf{F}_{ij}$ denotes the expected number of *new infections* entering the $i$–th infected compartment during $[t, t+1)$ that are generated by a single infectious individual in the $j$–th infected compartment at time $t$. By contrast, $\mathbf{T}_{ij}$ denotes the *fraction* (or equivalently, the probability per time step) of individuals in the $j$–th infected compartment who, without generating new infections, remain infectious and move to the $i$–th infected compartment over the time interval $[t, t+1)$. In line with this interpretation of the entry of $\mathbf{T}$, the sums of the columns of $\mathbf{T}$ should always be less than or equal to 1 [11]. The next–generation matrix is given by

$$\mathbf{NGM} = \mathbf{F}(\mathbf{I} - \mathbf{T})^{-1},$$

where $\mathbf{I}$ is the identity matrix. The $(i, j)$ entry of the matrix $(\mathbf{I} - \mathbf{T})^{-1}$ can be interpreted as the average amount of time that an infected individual introduced into compartment $j$ spends in compartment $i$ over its infectious lifetime. As for the epidemiological interpretation of the **NGM** matrix, its $(i, j)$ entry represents the expected number of secondary cases in compartment $i$ produced by a single infectious individual in compartment $j$ [10]. The basic reproduction number $\mathcal{R}_0$ (or $\mathcal{R}_c$, if containment measures are in place) is the spectral radius of **NGM**.

We therefore begin by computing the disease–free equilibrium of system (3), that is

$$E_0 = (S_1^0,\, V_1^0,\, 0,\, 0,\, S_2^0,\, V_2^0,\, 0,\, 0),$$

where

$$S_1^0 = \frac{(1-p)\Lambda}{A_1}, \qquad S_2^0 = \frac{r_1\eta}{\Delta}\left\{\frac{(1-p)\Lambda A_2 B_2 + r_2\omega[A_1 p\Lambda + r_1\psi_1(1-p)\Lambda]}{A_1 A_2}\right\},$$
$$V_1^0 = \frac{A_1 p\Lambda + r_1\psi_1(1-p)\Lambda}{A_1 A_2}, \quad V_2^0 = \frac{r_1\eta}{\Delta}\left[\frac{A_1 B_1 p\Lambda + B_1 r_1\psi_1(1-p)\Lambda + A_2 r_2\psi_2(1-p)\Lambda}{A_1 A_2}\right],$$

with $A_1 = 1 - r_1(1-\psi_1-\eta)$, $A_2 = 1 - r_1(1-\eta)$, $B_1 = 1 - r_2(1-\psi_2)$, $B_2 = 1 - r_2(1-\omega)$ and $\Delta = (1-r_2)[1 - r_2(1-\psi_2-\omega)]$. In fact, the disease–free equilibrium coordinates are the solutions of the following linear–affine system:

$$S_1^0 = (1-p)\Lambda + r_1\left[1-\psi_1-\eta\right] S_1^0, \qquad V_1^0 = p\Lambda + r_1(1-\eta)\, V_1^0 + r_1\psi_1\, S_1^0,$$
$$S_2^0 = r_1\eta\, S_1^0 + r_2(1-\psi_2)\, S_2^0 + r_2\omega\, V_2^0, \qquad V_2^0 = r_1\eta\, V_1^0 + r_2\psi_2\, S_2^0 + r_2(1-\omega)\, V_2^0.$$

The equations on the first line give the expressions of $S_1^0$ and $V_1^0$. As for the equations on the second line, they can be rewritten in matrix form as

$$\mathbf{M}\begin{pmatrix} S_2^0 \\ V_2^0 \end{pmatrix} = \begin{pmatrix} r_1\eta\, S_1^0 \\ r_1\eta\, V_1^0 \end{pmatrix}, \quad \text{with} \quad \mathbf{M} = \begin{pmatrix} B_1 & -r_2\omega \\ -r_2\psi_2 & B_2 \end{pmatrix}.$$

Therefore, provided that

$$\Delta := \det(\mathbf{M}) = B_1 B_2 - r_2^2\omega\psi_2 = (1-r_2)\big[1 - r_2(1-\psi_2-\omega)\big] \neq 0,$$

the inversion of $\mathbf{M}$ yields the coordinates $S_2^0$ and $V_2^0$,

$$S_2^0 = \frac{r_1\eta}{\Delta}\Big(B_2\, S_1^0 + r_2\omega\, V_1^0\Big), \qquad V_2^0 = \frac{r_1\eta}{\Delta}\Big(r_2\psi_2\, S_1^0 + B_1 V_1^0\Big).$$

To derive the expression of the control reproduction number $\mathcal{R}_c$, we first decompose the infected subsystem into $\mathcal{F}(\mathbf{x}(t))$ and $\mathcal{T}(\mathbf{x}(t))$. Note that $\mathcal{F}(\mathbf{x}(t))$ should include only infections that are newly arising, but does not include terms which describe the transfer of infectious individuals from one infected compartment to another [7]. In particular, the transfer terms of infectious individuals due to ageing from infants to adults belong to $\mathcal{T}(\mathbf{x}(t))$, not to $\mathcal{F}(\mathbf{x}(t))$. With this convention, the decomposition is

$$\mathcal{F}(\mathbf{x}(t)) = \begin{pmatrix} r_1 G_1(t)[S_1(t) + \sigma_1 V_1(t)] \\ r_2 G_2(t)[S_2(t) + \sigma_2 V_2(t)] \end{pmatrix}, \qquad \mathcal{T}(\mathbf{x}(t)) = \begin{pmatrix} r_3(1-\gamma_1-\eta) I_1(t) \\ r_3\eta I_1(t) + r_2(1-\gamma_2) I_2(t) \end{pmatrix},$$

then we compute the fertility and transition matrices $\mathbf{F}$ and $\mathbf{T}$:

$$\mathbf{F} = \begin{pmatrix} C_1 g_{11} & C_1 g_{12} \\ C_2 g_{21} & C_2 g_{22} \end{pmatrix}, \qquad \mathbf{T} = \begin{pmatrix} r_3(1-\gamma_1-\eta) & 0 \\ r_3\eta & r_2(1-\gamma_2) \end{pmatrix},$$

where

$$C_i := r_i\big[S_i^0 + \sigma_i V_i^0\big] \quad \text{and} \quad g_{ij} := \left.\frac{\partial G_i}{\partial I_j}\right|_{E_0} = \frac{\beta_{ij}}{N_j^0}, \qquad i,j = 1,2.$$

The next–generation matrix is then

$$\mathbf{NGM} = \begin{pmatrix} \dfrac{C_1 g_{11}}{a} + \dfrac{C_1 g_{12}\, r_3\eta}{ab} & \dfrac{C_1 g_{12}}{b} \\ \dfrac{C_2 g_{21}}{a} + \dfrac{C_2 g_{22}\, r_3\eta}{ab} & \dfrac{C_2 g_{22}}{b} \end{pmatrix},$$

where

$$a = 1 - r_3(1-\gamma_1-\eta), \qquad b = 1 - r_2(1-\gamma_2).$$

Note that each entry in the first column of **NGM** is the sum of two terms. This is consistent with the epidemiological meaning of the entries of **NGM**. In particular, $\mathbf{NGM}_{11}$ is the expected number of infected infants generated by a single infant index case over its infectious period. For an infant index case, the term $C_1 g_{11}/a$ indicates the number of infections produced before ageing, whereas $C_1 g_{12} r_3 \eta/(ab)$ accounts for the infections generated after ageing. Similarly, $\mathbf{NGM}_{21}$ is the number of infected adults produced by an infant index case: the term $C_2 g_{21}/a$ counts infections *before* ageing, and $C_2 g_{22} r_3 \eta/(ab)$ counts infections *after* ageing.

The control reproduction number $\mathcal{R}_c$ is the spectral radius of NGM, that is

$$\mathcal{R}_c = \frac{\operatorname{tr}\mathbf{NGM} + \sqrt{(\operatorname{tr}\mathbf{NGM})^2 - 4\det\mathbf{NGM}}}{2},$$

with

$$\operatorname{tr}\mathbf{NGM} = \frac{C_1 g_{11}}{a} + \frac{C_1 g_{12}\, r_3 \eta}{ab} + \frac{C_2 g_{22}}{b}, \qquad \det\mathbf{NGM} = \frac{C_1 C_2}{ab}\big(g_{11}g_{22} - g_{12}g_{21}\big),$$

that is,

$$\begin{aligned}\mathcal{R}_c = {}& \frac{C_1\, g_{11}}{2a} + \frac{C_1\, g_{12}\, r_3\, \eta}{2ab} + \frac{C_2\, g_{22}}{2b} \\ &+ \left[\left(\frac{C_1\, g_{11}}{2a} + \frac{C_1\, g_{12}\, r_3\, \eta}{2ab} + \frac{C_2\, g_{22}}{2b}\right)^2 - \frac{C_1 C_2}{ab}\big(g_{11}g_{22} - g_{12}g_{21}\big)\right]^{1/2}.\end{aligned} \tag{S.1}$$

In the absence of control strategies, the disease–free equilibrium of model (3) reduces to $E_0 = (S_1^0, 0, 0, 0, S_2^0, 0, 0, 0)$, where, assuming that $1 - r_1(1-\eta) \neq 0$ and $1 - r_2 \neq 0$,

$$S_1^0 = \frac{\Lambda}{1 - r_1(1-\eta)}, \quad S_2^0 = \frac{r_1 \eta\, \Lambda}{(1-r_2)[1 - r_1(1-\eta)]}.$$

The expression of the fertility matrix simplifies to

$$\mathbf{F} = \begin{pmatrix} r_1\beta_{11} & \dfrac{1-r_2}{\eta}\,\beta_{12} \\ \dfrac{r_1 r_2 \eta}{1-r_2}\,\beta_{21} & r_2\beta_{22} \end{pmatrix},$$

since $S_2^0/S_1^0 = r_1\eta/(1-r_2)$. The next–generation matrix becomes

$$\mathbf{NGM} = \begin{pmatrix} \dfrac{r_1\beta_{11}}{a} + \dfrac{(1-r_2) r_3 \beta_{12}}{ab} & \dfrac{1-r_2}{\eta b}\beta_{12} \\ \dfrac{r_1 r_2 \eta}{(1-r_2)a}\beta_{21} + \dfrac{r_2 r_3 \eta}{ab}\,\beta_{22} & \dfrac{r_2\beta_{22}}{b} \end{pmatrix}.$$

In this case

$$\operatorname{tr}\mathbf{NGM} = \frac{r_1\, \beta_{11}}{a} + \frac{(1-r_2)\, r_3\, \beta_{12}}{ab} + \frac{r_2\, \beta_{22}}{b}, \qquad \det\mathbf{NGM} = \frac{r_1 r_2}{ab}\big(\beta_{11}\beta_{22} - \beta_{12}\beta_{21}\big).$$

The basic reproduction number is then given by

$$
\begin{aligned}
\mathcal{R}_0 &= \frac{r_1\,\beta_{11}}{2a} + \frac{(1-r_2)\,r_3\,\beta_{12}}{2ab} + \frac{r_2\,\beta_{22}}{2b} \\
&\quad + \left[\left(\frac{r_1\,\beta_{11}}{2a} + \frac{(1-r_2)\,r_3\,\beta_{12}}{2ab} + \frac{r_2\,\beta_{22}}{2b}\right)^2 - \frac{r_1 r_2}{ab}\left(\beta_{11}\beta_{22} - \beta_{12}\beta_{21}\right)\right]^{1/2},
\end{aligned}
$$

where $a = 1 - r_3(1-\gamma_1-\eta)$ and $b = 1 - r_2(1-\gamma_2)$.

As mentioned in Section 3 of the main text, one of the most important properties of $\mathcal{R}_c$ (or $\mathcal{R}_0$, equivalently) is its role as an epidemic threshold: if $\mathcal{R}_c < 1$, then one infectious individual generates less than one secondary case and the disease is expected to die out, whereas if $\mathcal{R}_c > 1$, then the disease spreads in the population. This property is mathematically formalised by saying that the disease-free equilibrium is *locally asymptotically stable* when $\mathcal{R}_c < 1$. This means that when $\mathcal{R}_c < 1$, small introductions of infection (e.g., a few cases) tend to die out: the system naturally returns to a situation with no sustained transmission. In other words, as long as the outbreak starts small, it does not take off. The following proposition mathematically proves this property.

**Proposition 2.** If $\mathcal{R}_c < 1$, then the disease–free equilibrium $E_0$ of the model is locally asymptotically stable. If $\mathcal{R}_c > 1$, then $E_0$ is unstable.

*Proof.* The next–generation matrix method for discrete–time systems [1] states that $E_0$ is locally asymptotically stable when $\mathcal{R}_c < 1$, provided that the matrix $\mathbf{F}+\mathbf{T}$ is irreducible and that $E_0$ is locally asymptotically stable in the disease–free submanifold. The matrix $\mathbf{F}+\mathbf{T}$ is irreducible, since $\beta_{ij} > 0$ for all $i,j = 1,2$. Let us check that $E_0$ is locally asymptotically stable in the disease–free submanifold. In this case, the system reduces to the following linear–affine system:

$$
\begin{aligned}
S_1(t+1) &= (1-p)\Lambda + r_1 S_1(t) - r_1\psi_1 S_1(t) - r_1\eta S_1(t), \\
V_1(t+1) &= p\Lambda + r_1 V_1(t) + r_1\psi_1 S_1(t) - r_1\eta V_1(t), \\
R_1(t+1) &= r_1(1-\eta) R_1(t), \\
S_2(t+1) &= r_1\eta S_1(t) + r_2\nu R_2(t) + r_2 S_2(t) - r_2\psi_2 S_2(t) + r_2\omega V_2(t), \\
V_2(t+1) &= r_1\eta V_1(t) + r_2\psi_2 S_2(t) - r_2\omega V_2(t) + r_2 V_2(t), \\
R_2(t+1) &= r_1\eta R_1(t) + r_2(1-\nu) R_2(t),
\end{aligned}
$$

that can be written in matrix form as $\mathbf{x}(t+1) = \mathbf{A}\mathbf{x}(t) + \mathbf{b}$, where

$$
\mathbf{x} = (S_1, V_1, R_1, S_2, V_2, R_2)^t, \quad \mathbf{b} = \big((1-p)\Lambda,\ p\Lambda,\ 0,\ 0,0,0\big)^t
$$

and

$$
\mathbf{A} = \begin{pmatrix}
r_1(1-\psi_1-\eta) & 0 & 0 & 0 & 0 & 0 \\
r_1\psi_1 & r_1(1-\eta) & 0 & 0 & 0 & 0 \\
0 & 0 & r_1(1-\eta) & 0 & 0 & 0 \\
r_1\eta & 0 & 0 & r_2(1-\psi_2) & r_2\omega & r_2\nu \\
0 & r_1\eta & 0 & r_2\psi_2 & r_2(1-\omega) & 0 \\
0 & 0 & r_1\eta & 0 & 0 & r_2(1-\nu)
\end{pmatrix}.
$$

The first four eigenvalues of $\mathbf{A}$ are $\lambda_1 = r_1(1-\psi_1-\eta)$, $\lambda_2 = \lambda_3 = r_1(1-\eta)$, $\lambda_4 = r_2(1-\nu)$, while the other two are the eigenvalues of the sub-matrix $\mathbf{B} = r_2\tilde{\mathbf{B}}$, with

$$
\tilde{\mathbf{B}} = \begin{pmatrix} (1-\psi_2) & \omega \\ \psi_2 & (1-\omega) \end{pmatrix},
$$

which are the roots of the characteristic polynomial $\lambda^2 - [2-(\psi_2+\omega)]\lambda + [1-(\psi_2+\omega)]$, that can be factorized as $(\lambda-1)[\lambda-(1-\psi_2-\omega)]$. So the last two eigenvalues of $B$ are

$$\lambda_5 = r_2, \qquad \lambda_6 = r_2(1-\psi_2-\omega).$$

Since all the eigenvalues have modulus strictly less than 1, $\rho(\mathbf{A}) < 1$ and $E_0$ is locally asymptotically stable in the disease–free submanifold. Hence, $E_0$ is locally asymptotically stable for system (3). □
